# Five-Year Glycemic Trends in a Remote Diabetes Monitoring Program (RDMP): Contributions of Program Engagement and Evolving Pharmacotherapy Adoption and Adherence

**DOI:** 10.64898/2026.10.08.26363772

**Authors:** Roberta James, Meeraj Kothari, Ellen Almirol, Christa-Marie Singleton, George Verghese, Elizabeth Lowden

## Abstract

**Background:** Digital diabetes programs have expanded quickly, but evidence of durable long-term benefit remains limited. In addition, as diabetes treatment guidelines changed to include cardiorenal-protective antihyperglycemics as first-line therapy, the effects of their adoption on glycemic control and program engagement within these programs remain poorly understood.

**Objective:** To characterize longitudinal trends in glycemic control and estimate the extent to which these trends are associated with time-invariant and time-varying domains, among adults with T2D participating in Teladoc Health’s Remote Diabetes Monitoring Program (RDMP).

**Methods:** This was a retrospective, longitudinal cohort study of adults with T2D participating in the Teladoc RDMP from 2021-2025. Domains were split into time-invariant (i.e. demographics and baseline clinical characteristics) and time-varying covariates (e.g. antihyperglycemic treatment complexity/adherence, program utilization/engagement, and adoption of cardiorenal protective antihyperglycemics). Primary outcome was estimated A1c (eA1c), calculated from fasting blood glucose (FBG) measurements captured through cellular-enable smart glucose meters. Longitudinal mixed-effects models were used to estimate adjusted annual trends in eA1c over 5 years, and Gelbach decomposition to quantify statistical contributions across domains. Secondary analyses examined attainment of eA1c <7% using GEE and complementary decomposition methods.

**Results:** A total of 56,754 members with mean age 64.2 (SD 10.4) and 49% female were included in the analysis. Mean eA1c declined by 0.06 percentage points per year (95% CI −0.06 to −0.06) and remained significantly lower after full adjustment for demographic, antihyperglycemic treatment and cardiorenal-protective therapy adherence, and program engagement factors (β=−.04 percentage points/year; 95% CI −.04 to −.03), demonstrating durable glycemic improvement over the five-year study period. The full model attenuated 48.6% of the declining glycemic trend, indicating that glycemic improvements were only partially explained by demographic, treatment, and engagement factors. Gelbach decomposition attributed most of the difference between the base and fully adjusted trends to cardiorenal-protective therapies (170.8%). Program engagement factors contributed an offsetting share of the explained trend (−71.9%), although greater FBG monitoring and application engagement were associated with lower eA1c in adjusted analyses, while treatment history and complexity contributed minimally (1.1%). Adjusted odds of attaining eA1c <7% increased 8.0% annually (OR 1.08; 95% CI 1.07-1.09), corresponding to a 1.21-percentage-point annual increase in probability; adjusted predicted glycemic control increased from 75.2% in 2021 to 80.1% in 2025.

**Conclusion:** In this long-term experience with a large cohort of patients enrolled in a digital diabetes management program, progressive improvement in glycemic control was observed over the five years of follow-up. The pattern of glycemic improvement was consistent regardless of background exposure to GLP1-RAs and SGLT-2is therapies, including in participants who were never exposed to these therapies. These observations support guideline recommendations which include both the use of indicated medical therapies as well as lifestyle modification which can be facilitated with digital monitoring programs.

## INTRODUCTION

Digital diabetes programs have expanded quickly, but evidence of durable benefit remains limited. A 2024 assessment by the Peterson Health Technology Institute (PHTI) found that remote patient monitoring and behavioral lifestyle-modification programs produced small, short-term improvements in glycemic control.[1] A small number of multiyear studies of telemedicine and diabetes monitoring have been published, but high-quality long-term data remain limited for programs based strictly on digital monitoring.[2–4] Given the increasing use of these programs, their potential effects should be evaluated over longer follow-up periods. Additionally, the therapeutic landscape for type 2 diabetes (T2D) has shifted materially in recent years, potentially influencing observed secular trends. This study was designed to distinguish these concurrent influences. Durable improvement in glycemic control is necessary to reduce the risk of costly and serious complications.

Historically, pharmacologic management of T2D generally followed a stepwise treatment-intensification approach focused primarily on achieving glycemic targets, starting with lifestyle change prior to moving to metformin and eventually insulin for most uncontrolled patients. Trials of glucagon-like peptide-1 receptor agonists (GLP-1RAs) and sodium-glucose cotransporter-2 inhibitors (SGLT-2i) demonstrated risk reduction in costly diabetes-related comorbidities of atherosclerotic cardiovascular disease, heart failure, and renal disease. While the 2019 American Diabetes Association (ADA)/ European Association for the Study of Diabetes (EASD) consensus responded by recommending these agents based on cardiorenal risk, independent of HgbA1c[1,2], the 2022 update and additional society guidelines strengthened that position. [2,3] Tirzepatide, a dual glucose-dependent insulinotropic polypeptide (GIP/GLP-1RA) approved in 2022, has further expanded the therapeutic options available for improving glycemic control and weight-related outcomes in adults with T2D.[4] Between 2018 and 2022, prescribing shifted accordingly: SGLT-2 inhibitor use rose from 2.8% to 12.2%, and GLP-1RA use rose from 5.2% to 9.9%.[9] Additional studies have demonstrated a twofold increase in the adoption of GLP-1RAs and SGLT-2 inhibitors, although evidence regarding their effects on adherence and glycemic control in real-world settings remains limited. [10–12] Medication management alone, however, does not necessarily translate into sustained therapeutic exposure or durable glycemic benefit. In routine clinical practice, adherence may vary over time because of numerous patient- and treatment-related factors, many of which changed during this period. COVID-19 disrupted HbA1c testing, telehealth capabilities expanded, and shortages of GLP-1RA and incretin products occurred in 2022 and 2023, alongside formulary and step-therapy changes, out-of-pocket costs, and therapeutic switching.

The primary objective of this study is to characterize longitudinal trends in glycemic control among adults with T2D participating in the Teladoc Health’s RDMP from 2021 through 2025, adjusted for demographics and clinical characteristics, given secular changes over the last 5 years. This study aims to estimate the extent to which these trends are associated with time-varying antihyperglycemic treatment complexity, program utilization and engagement, and adoption and continued use of cardiorenal protective antihyperglycemics. Findings from this study may inform how future evaluations of digital diabetes programs account for treatment-era changes and identify program engagement and medication adherence patterns associated with sustained glycemic control that can guide program design.

## METHODS

### Teladoc Health Diabetes Management Program

The Teladoc Diabetes Program is a remote diabetes monitoring program (RDMP) designed to support self-management through connected technology and a web and mobile application that includes cellular-enabled smart blood glucose meter, unlimited testing supplies, personalized feedback, digital education, health information sharing with family and providers via a Health Summary report (HSR), and access to Certified Diabetes Care and Education Specialists (CDCES). Members of the program engage with coaches through scheduled one-on-one sessions, messaging, and outreach initiated in response to glucose excursions requiring additional support. This program complements usual clinical diabetes care that is generally managed by external providers.

### Study Design

This was a retrospective, longitudinal cohort study of adults with T2D participating in the Teladoc RDMP from 2021 through 2025. The primary unit of longitudinal analysis was at the member-period observation, with repeated observations nested within individual members.

Approval was granted by the Western Institutional Review Board (WIRB)-Copernicus Group (WCG) (IRB #20252125). All members provided consent to participate during enrollment into Teladoc Health Condition Management services, and guidelines outlined in the Declaration of Helsinki were followed. Participants were not compensated for their participation in the study.

### Population Selection

Members were eligible if they were 18 years of age at the start of the study period, had a self-reported diagnosis of T2D, were enrolled in the RDMP on or before January 1, 2021 and participated through December 31, 2025. Members also could not have evidence of continuous glucose monitor (CGM) use during the study period since glycemic control was measured based on fingerstick glucose (FBG) values used to estimate A1c per RDMP design. Members must have at least one prescription fill each year to identify antihyperglycemic medication utilization. Lastly, members were required to have at least one estimated A1c calculated from their records, creating a complete-five period panel per member.

### Data Source

Data were extracted from the Teladoc Health Registry, including: structured program engagement data, medication fill records, and FBG readings from the cellular-enabled capillary blood glucose meters. All study data were stored in Health Insurance Portability and Accountability Act (HIPAA)–compliant secure servers and were deidentified prior to analysis.

### Study Measurements

Study measures were organized into time-invariant baseline demographics and clinical characteristics and three time-varying domains: (1) antihyperglycemic treatment complexity and legacy antihyperglycemic medication management, (2) program utilization and engagement, and (3) cardiorenal protective antihyperglycemics (i.e.,I’s GLP-1RAs, dual GIP/GLP-1RAs, and SGLT-2is) adoption and adherence that shifted to ADA/AACE guideline first-line therapy during the study period.

#### Baseline Demographic and Clinical Characteristics

Demographic and clinical characteristics included age, sex, body mass index (BMI), self-reported HbA1c at baseline, hypertension status, duration of diabetes at program enrollment, and duration of participation in the RDMP (calculated) before the start of the study period.

Baseline treatment history characterized utilization of antihyperglycemic medication classes documented before the beginning of the observation period including prior use of GLP-1RAs, dual GIP/GLP-1RAs, SGLT-2is, Dipeptidyl peptidase-4 (DPP)-4 inhibitors, sulfonylureas, thiazolidinediones, meglitinides, biguanides, combination drugs (i.e. a class of drugs where multiple therapeutic classes are combined into a single dose), and insulin use.

Calendar year and time since program enrollment was included as distinct time measures. Calendar year was characterized as secular changes occurring from 2021 through 2025, whereas duration on the RDMP will account for differences in program tenure among observations occurring during the same calendar period. Time since enrollment was modeled continuously, with nonlinear specifications considered if supported by the data.

#### Antihyperglycemic Treatment Complexity and Legacy Medication Adherence

Time-varying treatment complexity served as a proxy for diabetes progression by the number and combination of active antihyperglycemic medication classes with and without insulin therapy and was measured for the previous year, i.e. treatment complexity reflected in 2021 was measured using 2020 prescription fills. A range from low to high diabetes complexity was defined as: no antihyperglycemic medications indicated low diabetes complexity, followed by 1 class of antihyperglycemic medication excluding insulin, 2 or more classes of antihyperglycemic without insulin, 2 or more classes of antihyperglycemic with one class as insulin, and then exclusively only insulin as the highest indicator of diabetes treatment complexity.

Where sufficient medication fill data were available, adherence to non-insulin antihyperglycemic therapies was characterized using proportion of days covered (PDC) as adherent (PDC≥0.8), nonadherent (PDC>0 and <0.8), and no prescription fill (PDC=0). Adherence measures were calculated separately by medication class of DPP-4i, sulfonylureas, thiazolidinediones, biguanides, and combination drugs and treated as time-varying measures during each reflecting medication availability during the relevant follow-up period. Because PDC cannot be calculated accurately for insulin, insulin utilization was coded as ‘Yes’ (1) if there is at least one prescription fill during each year period.

#### Program Utilization and Engagement

Program utilization and engagement were characterized by using structured program activity data summarized for each study year. Measures include the proportion of days with at least one FBG check, proportion of months with at least one web/mobile application login, at least one coaching interaction during the year, and sharing of the Health Summary Report. Coaching interactions varied between personalized health coaching sessions and alert-driven coaching outreach, prompted by glucose values on the cellular BGM that were above or below defined thresholds. Program engagement measures were interpreted as markers of observed utilization of the program rather than direct measures of intervention exposure or effectiveness.

#### Cardiorenal Protective Antihyperglycemics Adherence

Because of substantial changes in diabetes pharmacotherapy during the study period, utilization of three medication classes was characterized as distinct categories: GLP-1RAs; dual GIP/GLP-1RAs; and SGLT-2is. For analyses, using the previously defined adherence categories, medication exposure was also classified as adherent, non-adherent, and no prescription fill, using the same definition for the legacy antihyperglycemic medication classes.

This classification allowed medication to reflect both adoption and medication availability over time while retaining members without documented medication use as a clinically meaningful treatment state.

### Study Outcomes

The primary outcome was estimated A1c (eA1c), calculated from FBG measurements captured through the cellular-enabled smart glucose meter. For each eligible measurement window, FBG measurements are averaged over the preceding 90 days and are converted to an eA1c using the Nathan/ADAG equation[5]:

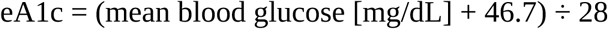

An estimate is eligible when it meets Teladoc’s observability and reporting framework within the same 90-day window. The observability framework evaluates whether the number and variability of a member’s FBG measurements are sufficient to support a reliable A1c estimate.[6] For each calendar year, the last eligible eA1c was selected as the annual outcome. This provided one annual glycemic outcome per member for longitudinal analyses. The secondary glycemic outcome was attainment of clinically relevant glycemic threshold, defined as eA1c <7.0% and measured each year. The threshold of 7.0% was selected given ADA recommended threshold for most adults with T2D; however, a person’s A1c goal is determined by their provider, as well as their age, disease and comorbidity complexity, life expectancy, cognitive function, and frequency or risk of hypoglycemic events.[7]

### Statistical Analysis

Baseline demographic and clinical characteristics were summarized for the overall longitudinal cohort and stratified by baseline self-reported A1c (<7% and ≥7%). Categorical variables were summarized using frequencies and percentages, and continuous variables using means and standard deviations (SDs). Time-varying treatment, medication adherence, and program engagement characteristics were summarized separately for each annual measurement period from 2021 through 2025.

The primary outcome analysis evaluated secular trends in continuous estimated A1c (eA1c) across the five annual measurement periods from 2021 through 2025 in the longitudinal closed-panel cohort. Linear mixed-effects models (LMMs) were used to account for repeated annual observations within members. Calendar year was modeled continuously to estimate the average annual change in eA1c during the study period. Models included a member-level random intercept to account for within-member correlation in repeated eA1c measurements. The coefficient for calendar year represents the estimated mean percentage-point change in eA1c per year and is reported with its 95% confidence interval (CI) and p-value. The unadjusted longitudinal model was specified as:

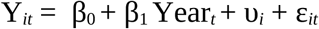

where *Y* _¿_is eA1c for member *i*during year *t*; *Yea r_t_* represents calendar year; *υ_i_*is a member-specific random intercept; and *ɛ* _¿_is the residual error. The coefficient *β*_1_represents the unadjusted mean annual change in eA1c.

To examine the degree to which the observed calendar-year trend was associated with differences in measured baseline characteristics and time-varying treatment and program factors, sequential models incorporated prespecified covariate blocks. Model 1 adjusted for time-invariant member demographic and baseline clinical characteristics 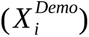. Model 2 additionally adjusted for time-varying treatment complexity, adherence to legacy antihyperglycemic medication classes, and insulin use 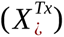. Model 3 further adjusted for measures of program engagement 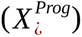. Model 4, the fully adjusted model, additionally incorporated adherence to cardiorenal protective antihyperglycemics (i.e. GLP-1 receptor agonists (GLP-1RAs), dual glucose-dependent insulinotropic polypeptide/GLP-1 receptor agonists (dual GIP/GLP-1RAs), and sodium-glucose cotransporter-2 inhibitors (SGLT-2is)) (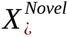). Thus, the fully adjusted was specified as:

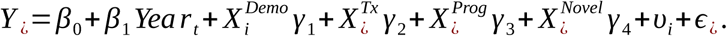

Each γ_k_ is that block’s coefficient vector. Each model was estimated three ways using identical analytic observations: (1) a linear mixed model (LMM) with a member-level random intercept and a random slope for year, with an unstructured covariance matrix for the random effects, fit by restricted maximum likelihood, which is the primary model for reporting the unadjusted and adjusted trend coefficients; (2) generalized estimating equations (GEE) with an identity link, exchangeable working correlation, and robust sandwich standard errors as a robustness analysis; and (3) pooled ordinary least squares (OLS) with member-clustered standard errors, which is used for the Gelbach decomposition because the omitted-variable-bias identity holds exactly only when the base and full models share the same estimator weights.

Because sequential addition of correlated covariates can make changes in the calendar-year coefficient dependent on the order in which covariates or domains enter the model, Gelbach’s decomposition was used to quantify the contribution of these three time-varying covariate domains to ethe difference between the demographic-adjusted (Model 1) and the fully adjusted calendar-year coefficients (Model 4).[8] The total explained attenuation is:

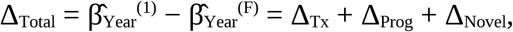

where β̂_Year_^(1)^ is the calendar-year coefficient from Model 1 and β̂_Year_^(F)^ is the corresponding coefficient from the fully adjusted Model 4, and where *Δ_Tx_*, *Δ_Prog_*, and *Δ_Novel_*represent the contributions of treatment, program engagement, and novel-therapy domains, respectively.

The primary decomposition was computed from pooled OLS. Standard errors and 95% confidence intervals for Δ_k_, Δ_Total_, and percentage contributions were obtained using a member-level nonparametric bootstrap with 2,000 replicates that resamples members and retains all repeated annual observations for each sampled member. Domain contributions were reported in eA1c percentage points per year and as percentages of the total change between the base and fully adjusted calendar-year coefficients.

A secondary analysis modeled the attainment of achieving an eA1c < 7% using the same four models and covariate domains but using GEE with a logit link function, exchangeable working correlation structure, and robust sandwich standard errors. The secondary trend was decomposed using a pooled linear probability model with member-clustered standard errors. Domain contribution 95% confidence intervals were obtained using the same member-level bootstrap method as the primary outcome analysis. The Karlson-Holm-Breen (KHB) method applied to a pooled logistic model with member-cluster standard errors was used as a robustness check on the logit scale[9].

Sensitivity analyses assessed the assumption of a constant linear temporal trend. First, calendar year was modeled categorically, with the first measurement year serving as the reference, allowing year-specific differences in mean eA1c to be estimated without imposing a linear trend. Second, the primary continuous-year analysis was repeated separately among members with baseline self-reported A1c ≥7% and <7% to evaluate whether longitudinal eA1c trends differed according to baseline glycemic status.

All hypothesis tests were two-sided, using an α=0.05 and estimates are reported with 95% confidence intervals. The primary analysis evaluated the linear calendar-year trend in mean eA1c. Gelbach domain contributions constituted a secondary family of statistical comparisons, with family-wise type I error controlled at 0.05 using the Holm procedure. Secondary outcome and sensitivity analyses were considered exploratory and hypothesis-generating; therefore, no adjustment for multiple comparisons was applied. All statistical analyses were conducted using R.4.4.3.

## RESULTS

Of the 621,070 members enrolled in the RDMP in 2021, 56,754 met all eligibility criteria (**Figure 1**). **Table 1** summarizes demographic and baseline clinical characteristics for the overall cohort and by self-reported A1c at enrollment: 34.8% had an A1c ≥7.0%, 45.5% had an A1c <7.0%, and 19.7% did not report an A1c. Overall, 50.0% of members were aged 65 years or older, and 49.3% were female. Mean program tenure at the start of the study was 1.0 year (SD, 0.7). Before 2021, 71% of members had used biguanides, 17.7% had used GLP-1RAs, 16.4% had used SGLT-2 inhibitors, and 17.1% had used insulin.

**Figure 1.**
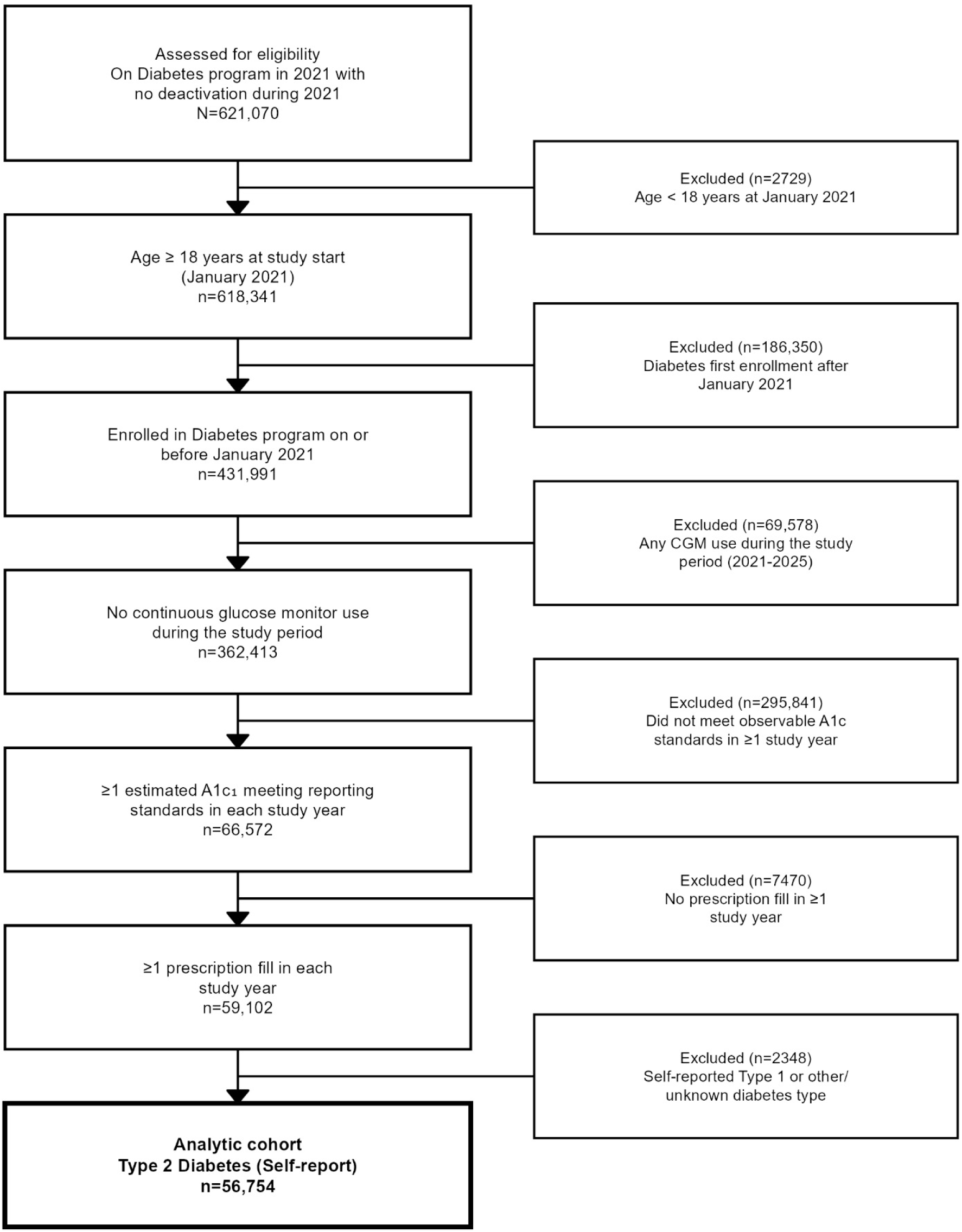
Analytic Cohort Flow Chart of Members with Type 2 Diabetes (T2D) enrolled in a Remote Diabetes Monitoring Program (RDMP) from January 2021 through December 2025.

**Table 1.**
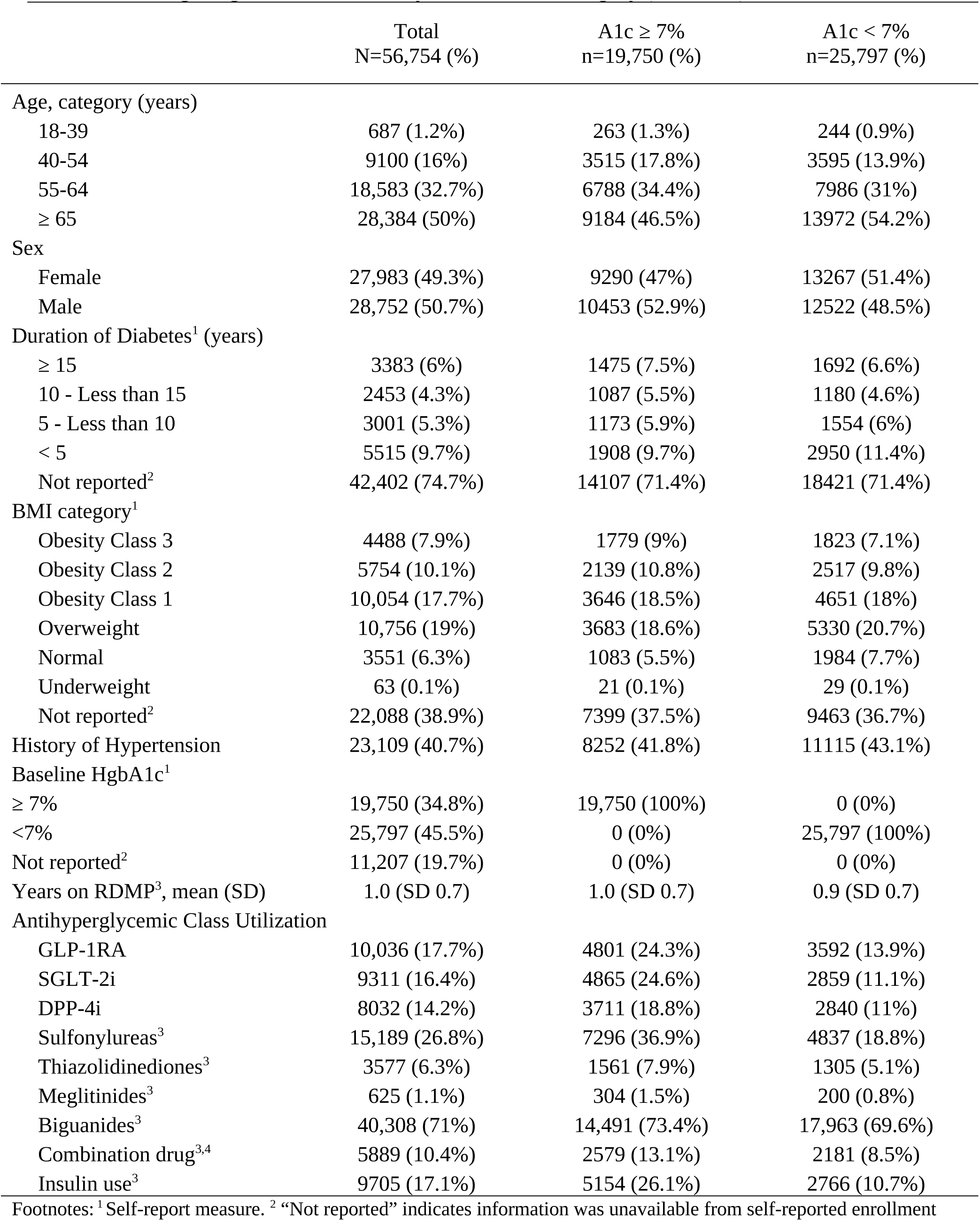
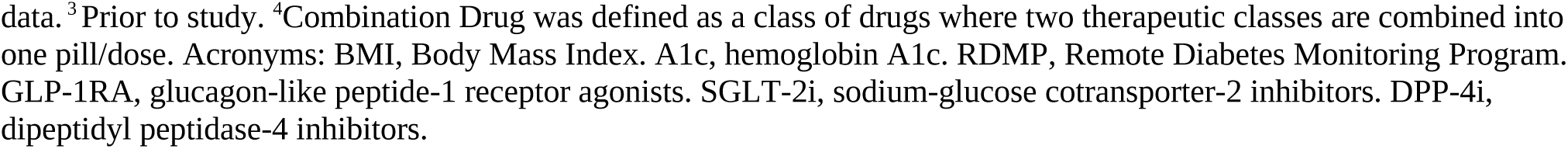
Baseline Demographic and Clinical Characteristics of Participants Enrolled in the Remote Diabetes Monitoring Program, Overall and by Baseline A1c Category (N=56,754)

**Table 2** displays the time-varying covariates by calendar year. Treatment complexity increased from 2021 to 2025 for the percentage of members on two or more classes of antihyperglycemics without insulin from 39.5% to 51.3%, while the percentage of members with only one class of antihyperglycemics decreased from 38.7% to 28.6%. The number of members not on antihyperglycemics decreased from 9.6% to 7.4%. Overall antihyperglycemic PDC remained stable over the five-year period at 0.8 (0.3). Members with any insulin use were stable during the study period at approximately 16%. Program utilization and engagement metrics remained mostly stable during the study period except sharing of the HSR that increased from mean counts of 0.2 (1.6) in 2021 to 1.8 (9.4) and 1.4 (5.6) in 2022 and 2023, respectively, and then decreased to 0.7 (3.3) and 0.6 (2.8) in 2024 and 2025, respectively. Among cardiorenal protective antihyperglycemics, GLP-1RA and dual GIP/GLP-1RA utilization increased from 22.1% in 2021 to 52.1% in 2025, while SGLT-2is increased from 18.6% to 32.7% over the study period. Biguanides had the largest decrease of almost 10% over five years.

**Table 2.**
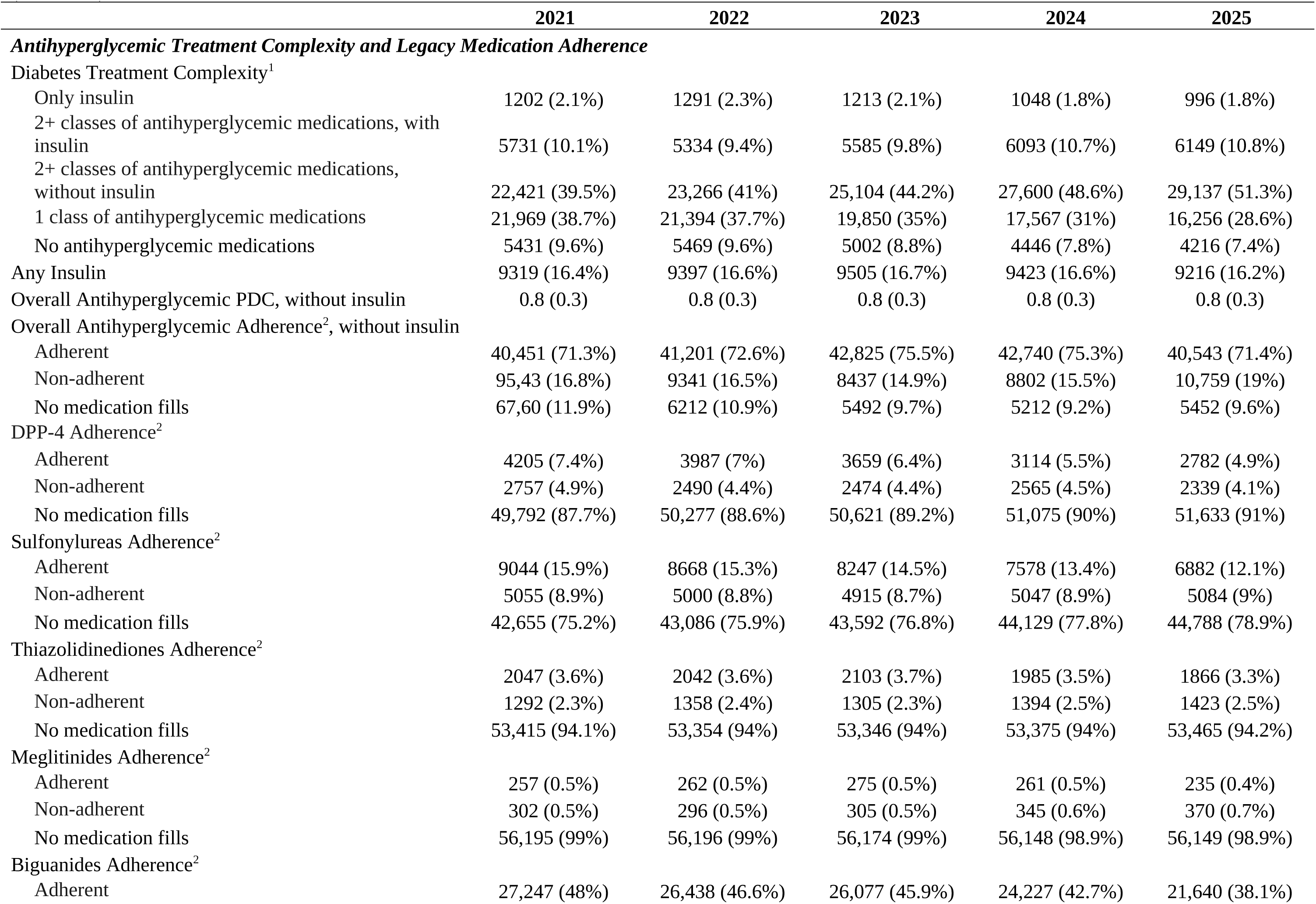
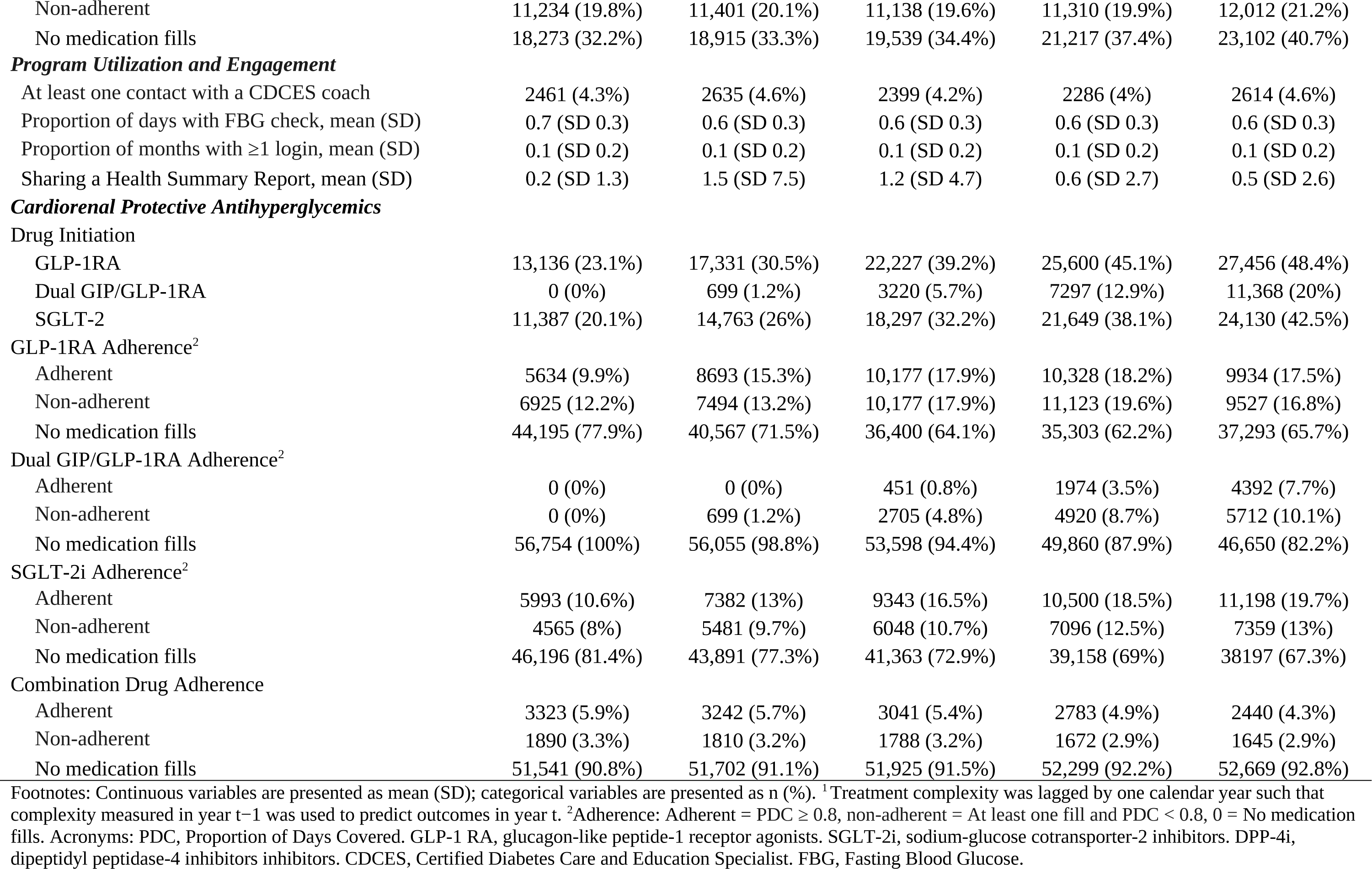
Annual Distribution of Time-Varying Covariates Among Participants Enrolled in the Remote Diabetes Monitoring Program, 2021-2025 (N=56,754)

**Figure 2** displays the durable reduction for the overall cohort and by self-reported A1c groups at enrollment. Unadjusted mean eA1c decreased from 6.5% in 2021 to 6.3% in 2025 among those whose mean self-reported A1c was 7.0% at the time of enrollment. **Figures 3a and b** display the utilization of the antihyperglycemic drug classes by year and the percentage of members utilizing the medication class who are adherent during the 12-month period, which also reflects availability of the drug especially for the GLP-1RAs and the new dual GIP/GLP-1RAs. **Figure 4** displays the unadjusted mean eA1c by those who never had a GLP-1RA or dual GIP/GLP-1RA, members who were on GLP-1RAs prior to 2021 and remained on GLP-1RAs, members who initiated GLP-1RAs or dual GIP/GLP-1RAs only in the follow up period, and members, and members who were not on GLP-1RAs at baseline and initiated SGLT-2is in the follow-up period. Members who received GLP-1RAs or dual GIP/GLP-1RAs has higher self-reported A1cs at enrollment ranging from 7.0% to 7.4% compared to 6.7% for those never receiving those drugs, likely indicating that people with higher-risk are being prioritized for the drug. All categories demonstrated reduction in eA1c by 2021 and were durable through 2025.

**Figure 2.**
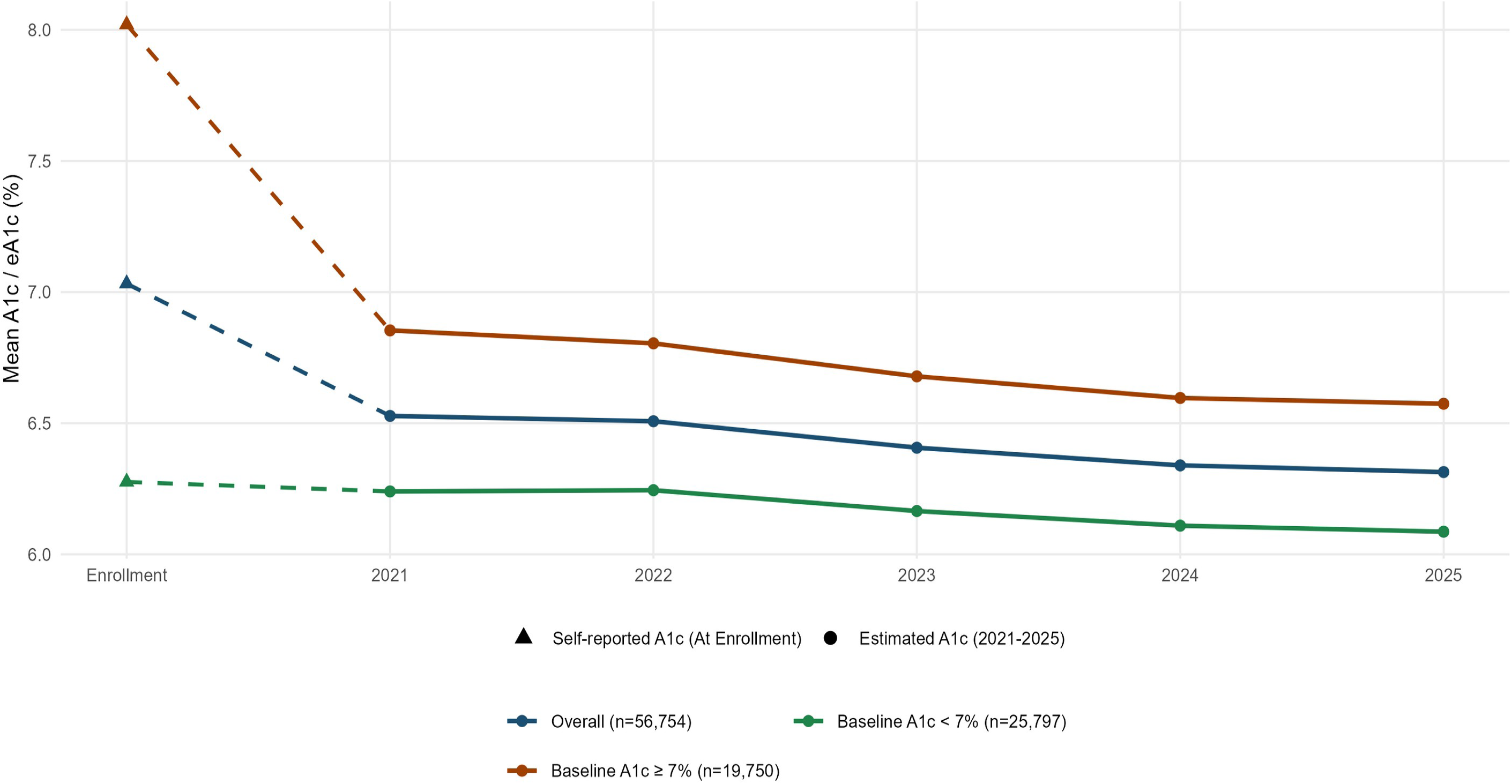
Mean Glycemic Control at Enrollment and Annual Follow-up (Estimated A1c), 2021-2025. Footnote: Dashed lines represent self-reported A1c at program enrollment. Solid lines and markers represent annual estimated A1c (eA1c) values. Error bars denote standard errors.

**Figure 3a.**
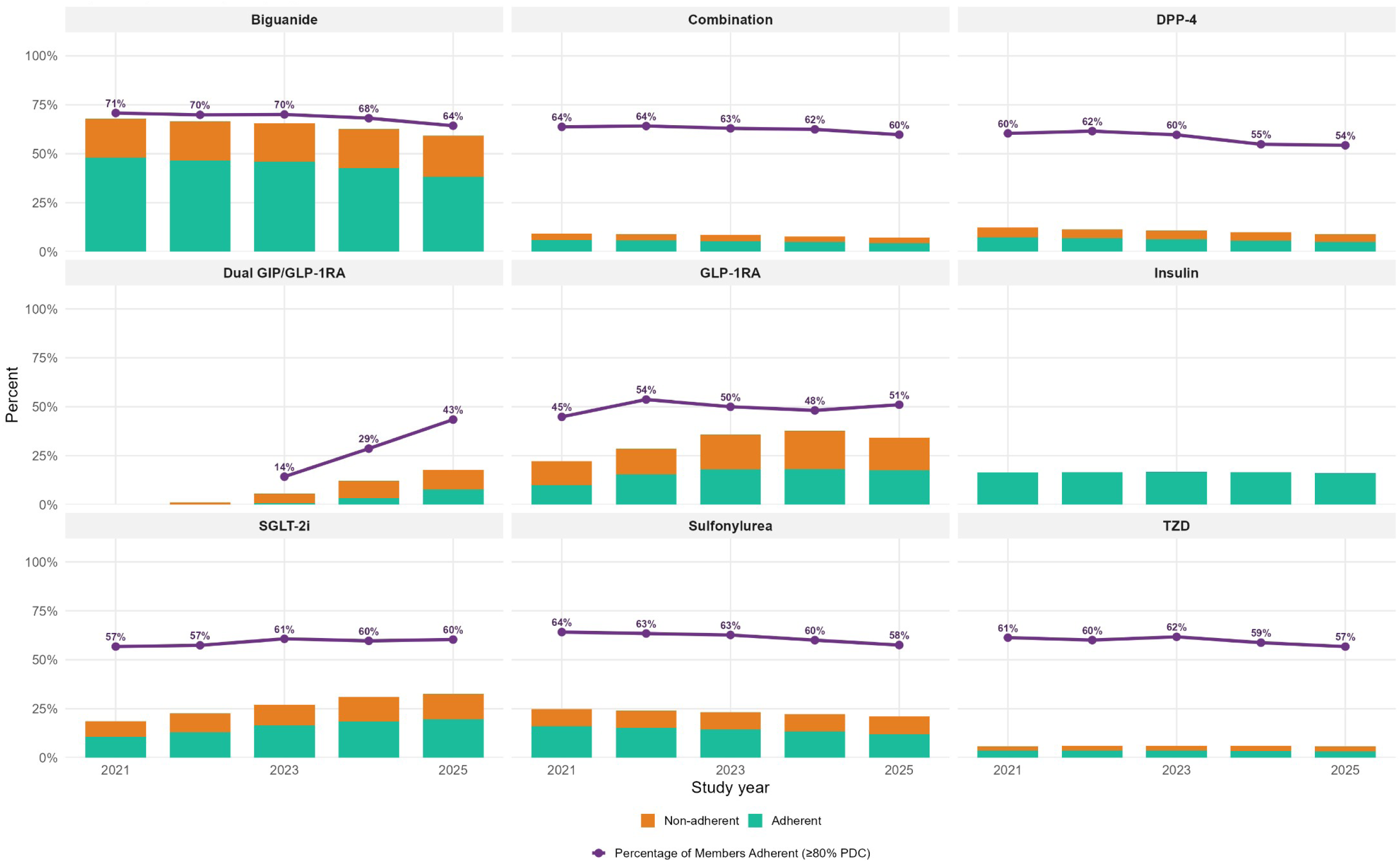
Annual Adherence to Antihyperglycemic Medication Classes, 2021-2025. Footnote: Green bars indicate adherent users (PDC ≥0.80), orange bars indicate non-adherent users (PDC <0.80 among users with ≥1 fill), and the purple line indicates overall utilization of each medication class. Insulin indicates adoption only. Meglitinides were excluded in this figure. Acronyms: DPP-4 = dipeptidyl peptidase-4 inhibitor; GIP = glucose-dependent insulinotropic polypeptide; GLP-1RA = glucagon-like peptide-1 receptor agonist; SGLT-2i = sodium-glucose cotransporter-2 inhibitor; TZD = thiazolidinedione. Combination = fixed-dose antihyperglycemic products.

**Figure 3b.**
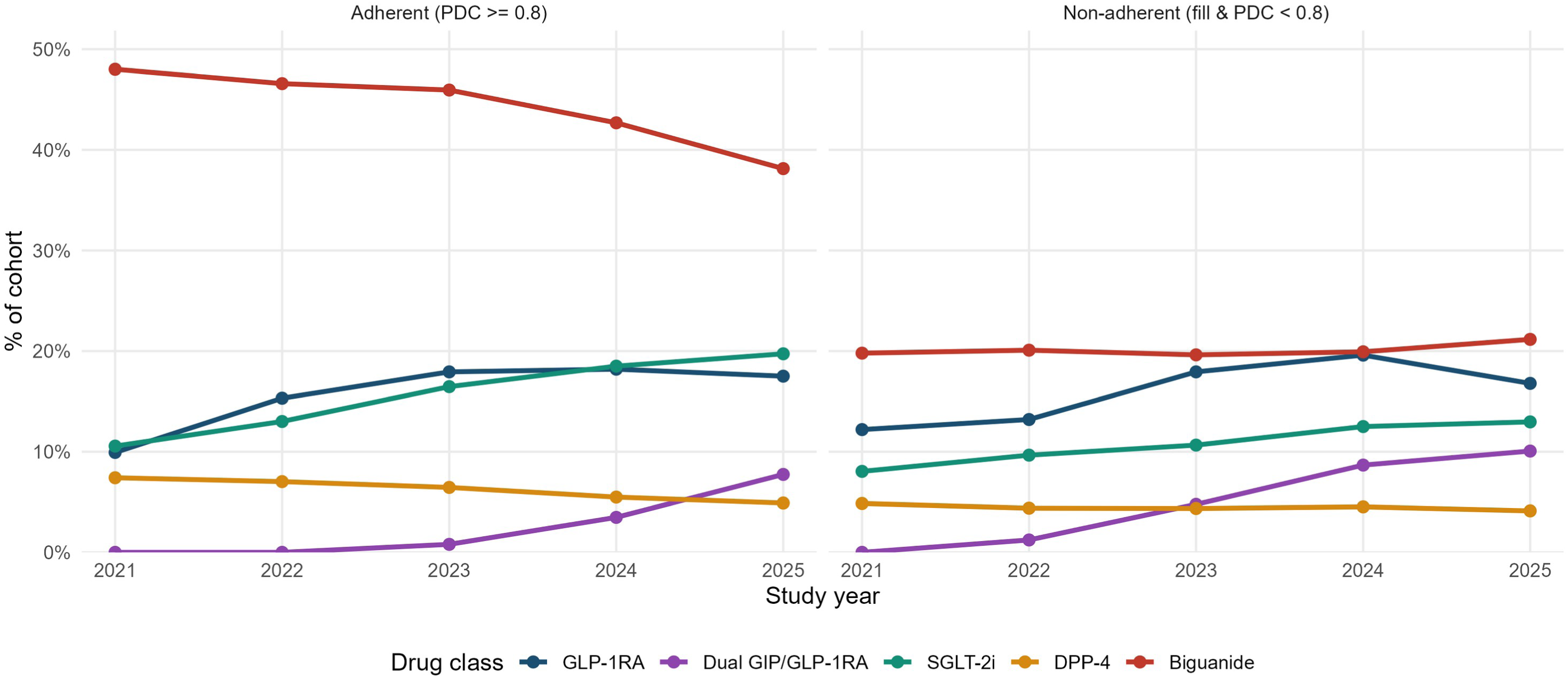
Distribution of Antihyperglycemic Medication Use1 and Adherence2 Status by Calendar Year. Footnote: ^1^Members with no medication fills during the study period were excluded from this figure. ^2^Adherence: Adherent was defined as PDC ≥0.8; Non-adherent was defined as at least one fill and PDC >0 and <0.8. Acronyms: PDC, Proportion of Days Covered. GLP-1RA, glucagon-like peptide-1 receptor agonists. Dual GIP/GLP-1RA, glucose-dependent insulinotropic polypeptide. SGLT-2i, sodium-glucose cotransporter-2 inhibitors. DPP-4i, dipeptidyl peptidase-4 inhibitors. JMIR Diabetes

**Figure 4.**
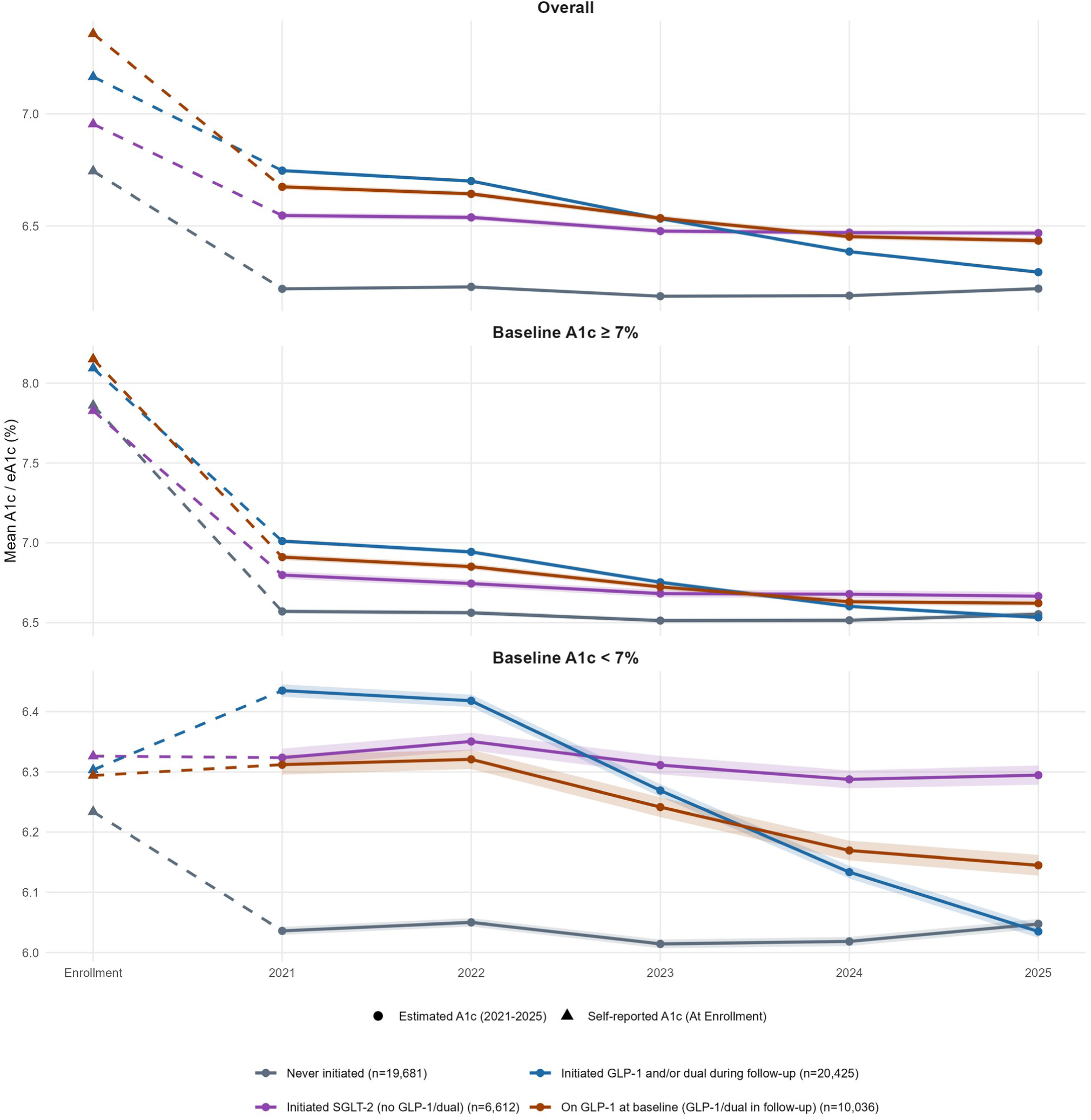
Unadjusted Mean Estimated A1c by Initiation of Cardiorenal-Protective Antihyperglycemic Therapies, 2021-2025. Footnotes: ^1^Treatment groups were mutually exclusive: (1) baseline GLP-1RA users who subsequently used GLP-1RA or dual GIP/GLP-1RA therapy (blue), (2) newly initiated GLP-1RA and/or dual GIP/GLP-1RA therapy during follow-up, (3) newly initiated SGLT-2 inhibitor therapy only (purple), and (4) never initiated any cardiorenal-protective antihyperglycemic therapy (gray). Dashed lines indicate from enrollment to 2021 (self-report A1c) and solid dot/lines indicate last observable Nathan (estimated A1c). Shaded bands represent standard errors.

### Primary Outcome Analysis

**Table 3** provides the unadjusted LMM results and the adjusted models with the time-varying domains. In the unadjusted LMM model, mean eA1c declined by 0.06 percentage points per year (95% CI, −0.06, −0.06). The annual decline remained after adjustment for demographic and baseline clinical characteristics (β= −.07; 95% CI; −0.07, −0.07), treatment complexity and legacy antihyperglycemic adherence (β= −.07; 95% CI; −0.71, −0.075), program utilization and engagement (β= −.06; 95% CI; −0.07, −0.06). In the fully adjusted model, which accounted for the cardiorenal protective antihyperglycemics, mean eA1c declined by 0.04 percentage points per year (95% CI; −0.04, −0.03). Proportion of days with FBG checks, proportion of months with web/mobile login, and sharing of the HSR were associated with lower eA1c, (β= −.05; 95% CI; −0.06, −0.03), (β= −.06; 95% CI; −0.87, −0.04), and (β= −.001; 95% CI; −0.002, −0.000), respectively, though sharing of the HSR was a very weak association. Coaching contact was associated with higher eA1c (β= .03; 95% CI; 0.01, 0.05), which likely indicates that members engaging with coaching had higher needs. The cardiorenal protective antihyperglycemics were all associated with lower eA1c with the dual GIP/GLP-1RAs adherence demonstrating the strongest association (β= −.50; 95% CI; −0.54, −0.46). The fully adjusted model accounted for 48.6% of the decline in eA1c trend and provided the best fit among the nested mixed-effects model with an Akaike information criterion of 326,709.1 compared with the 328,832.8 for the unadjusted model. Because the sequential changes in the year coefficient depend on the order in which correlated covariate domains enter the model, the prespecified Gelbach decomposition was used to quantify the contribution of each domain independently of entry order.

**Table 3.**
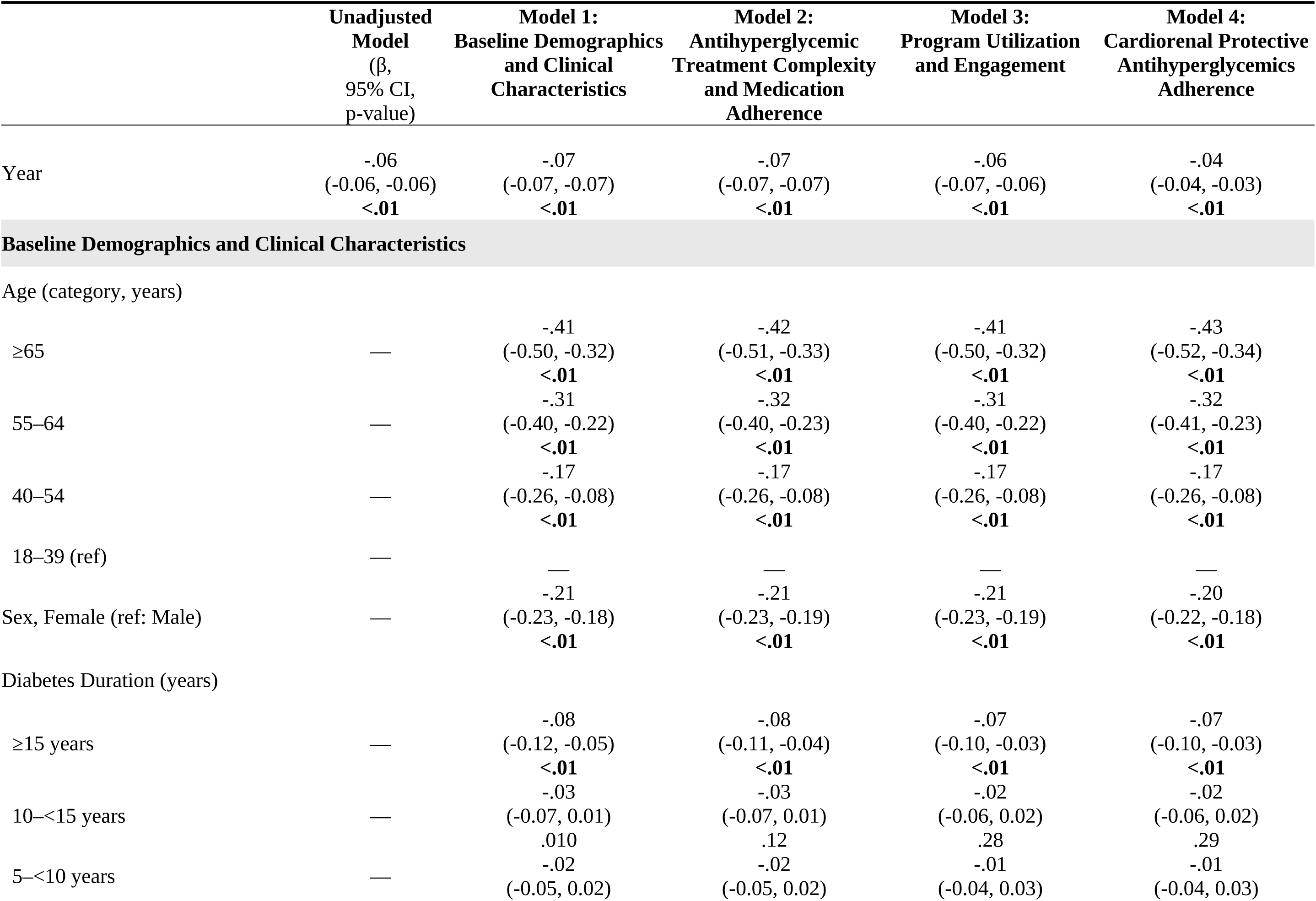
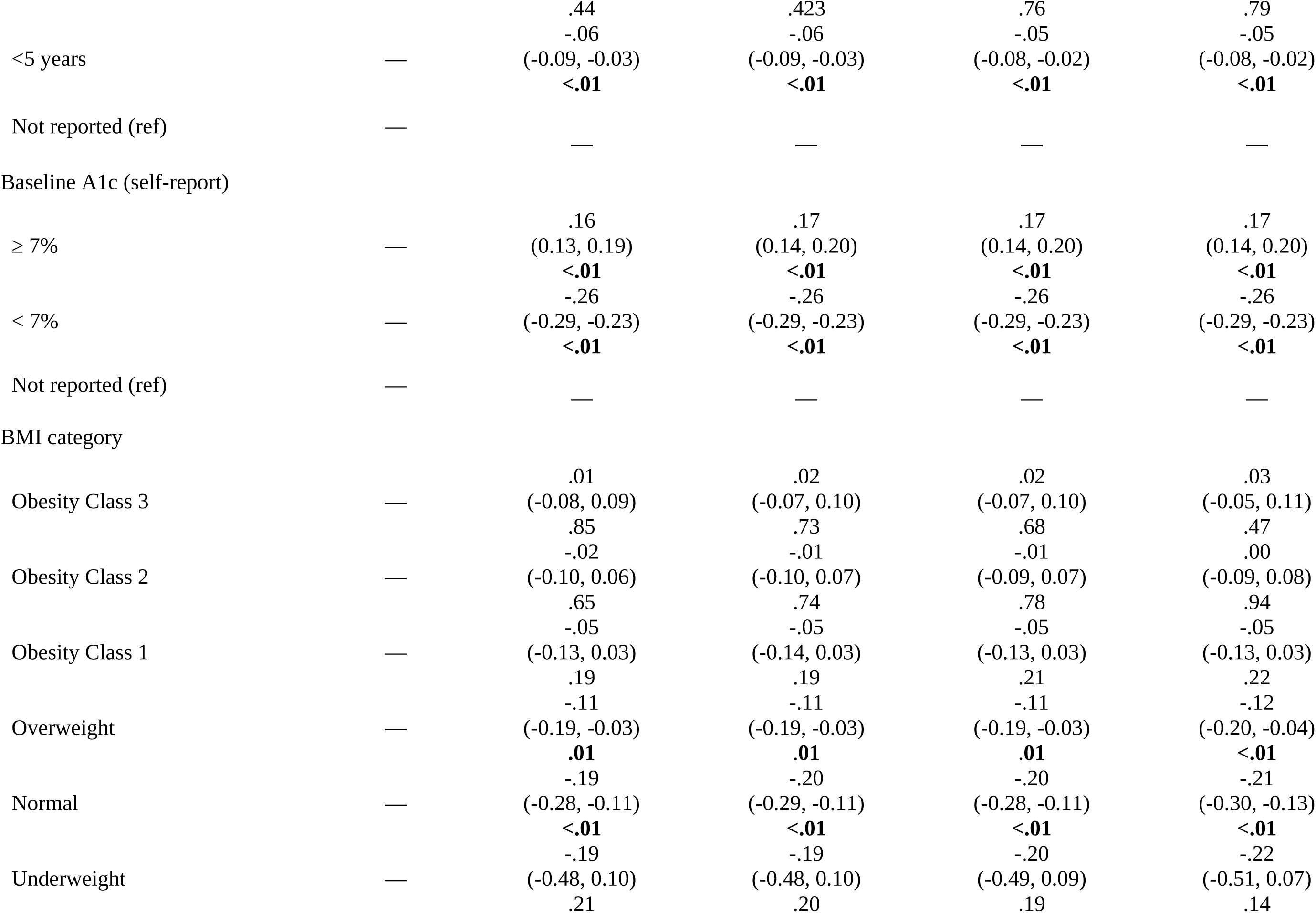
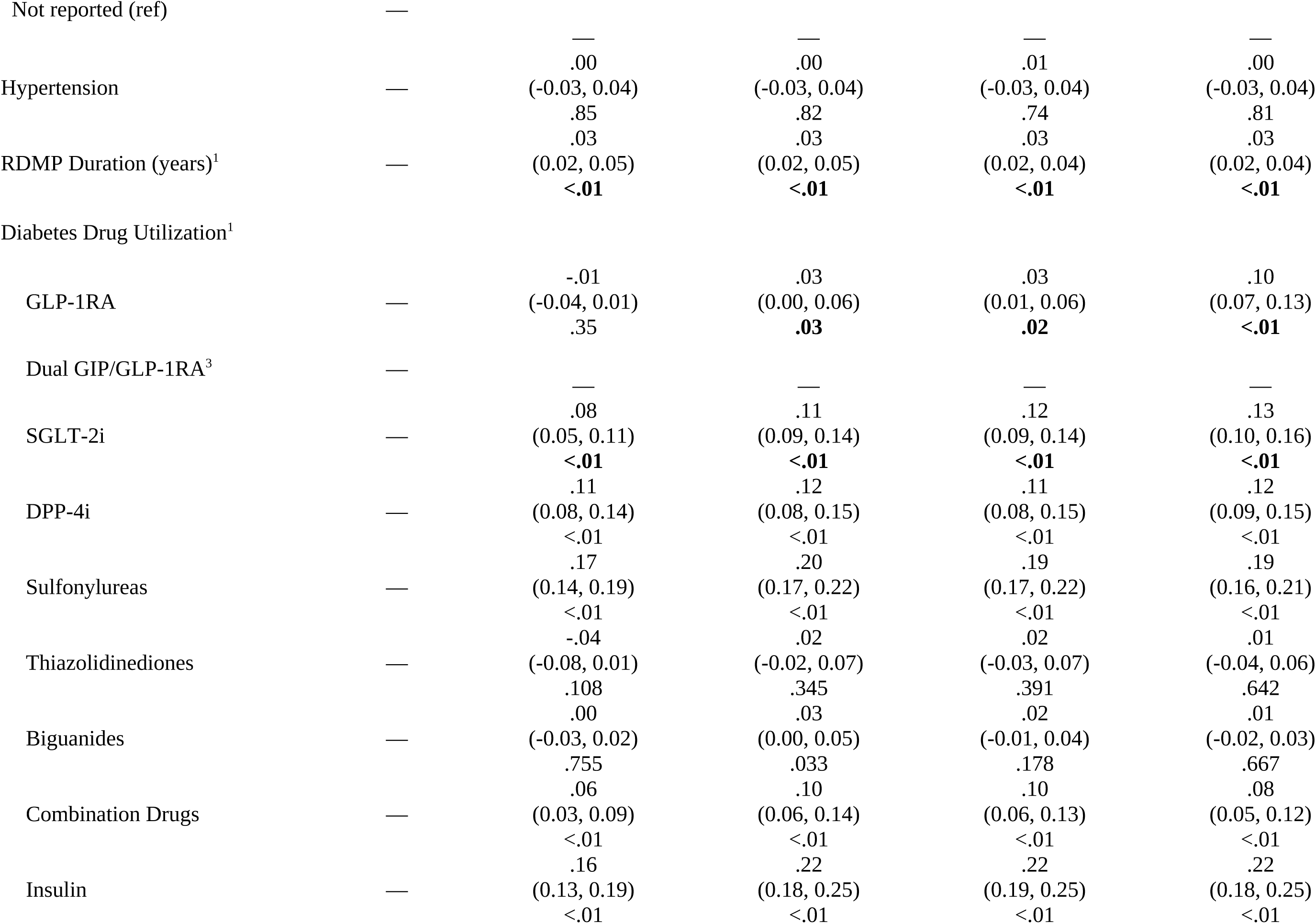
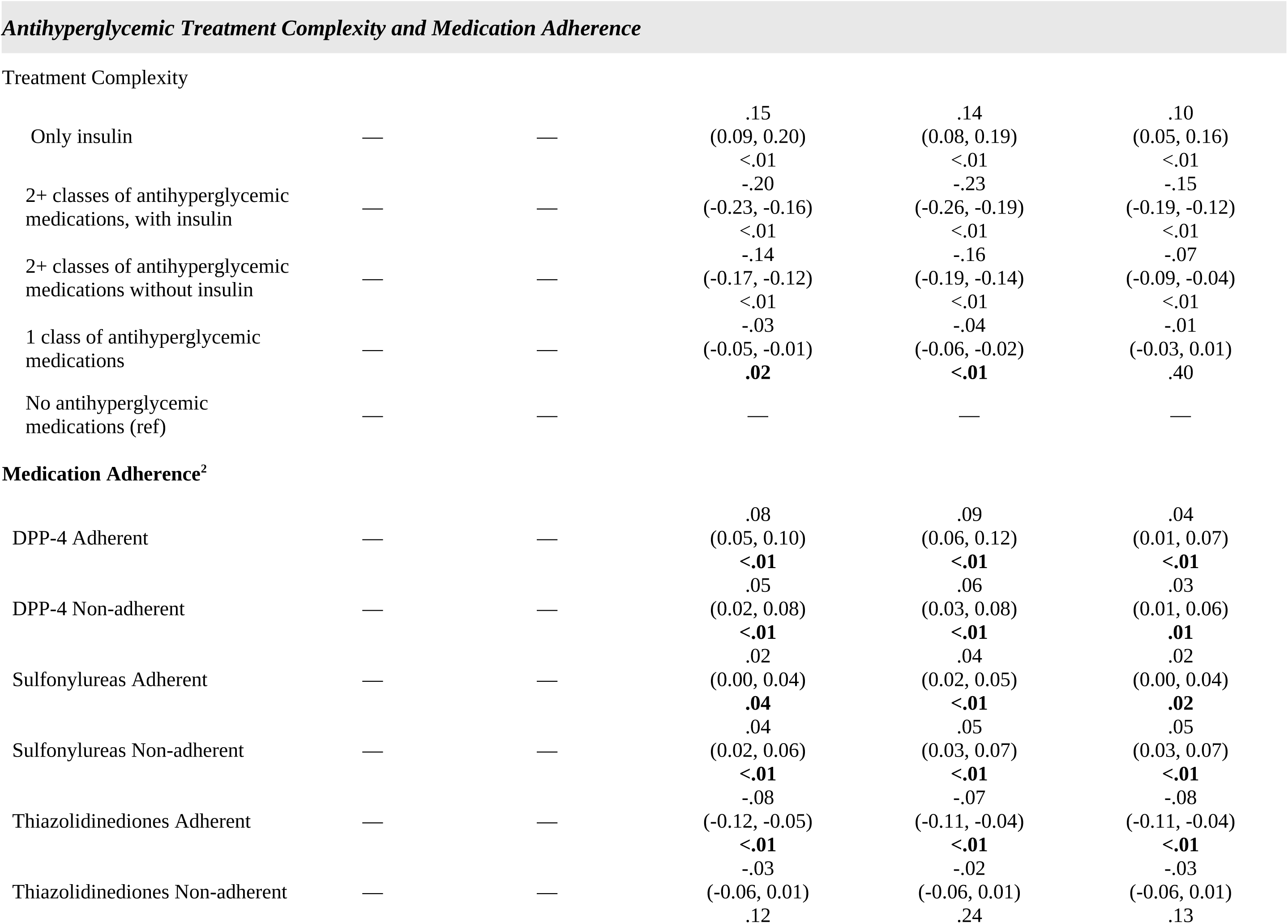
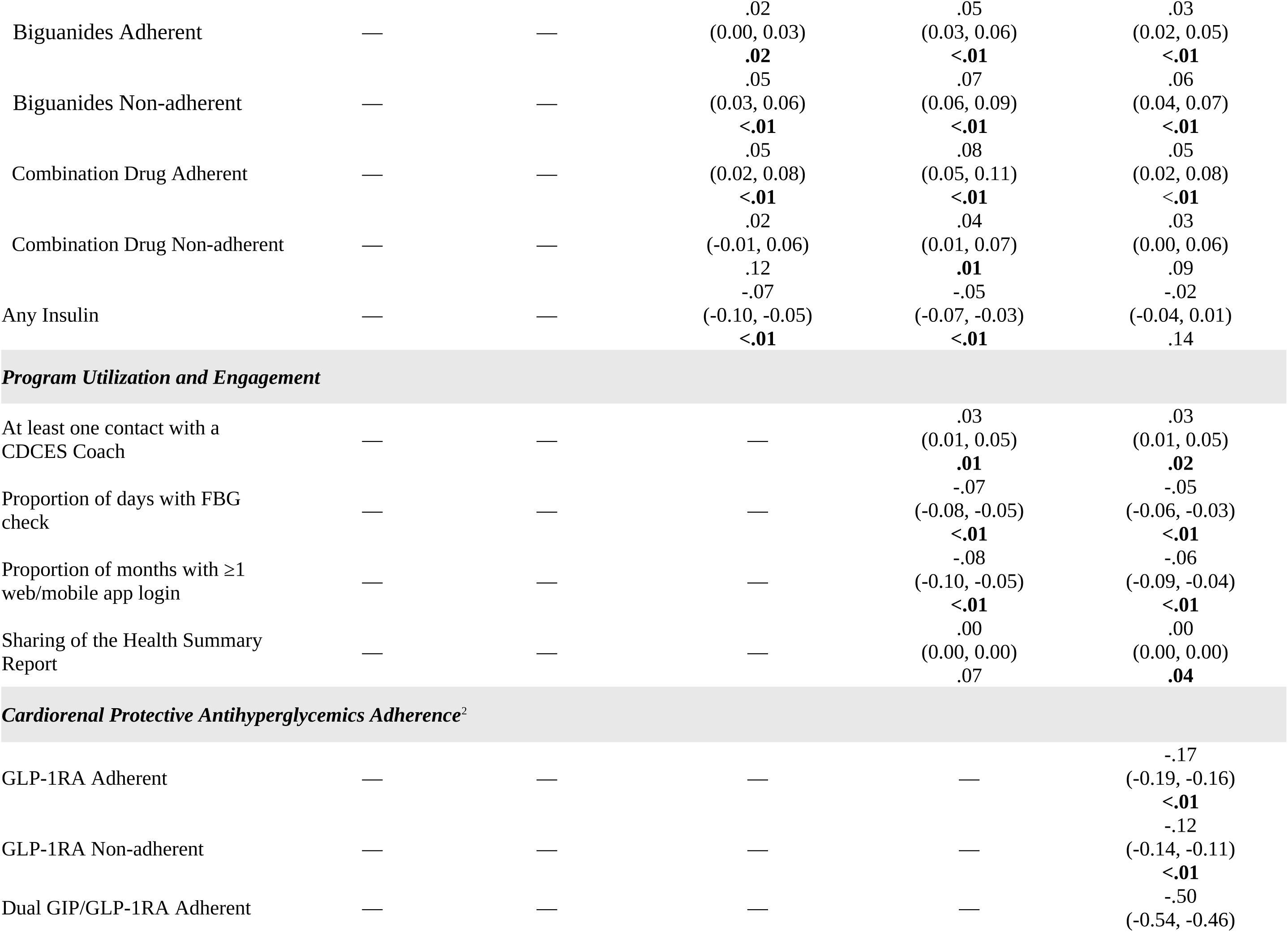
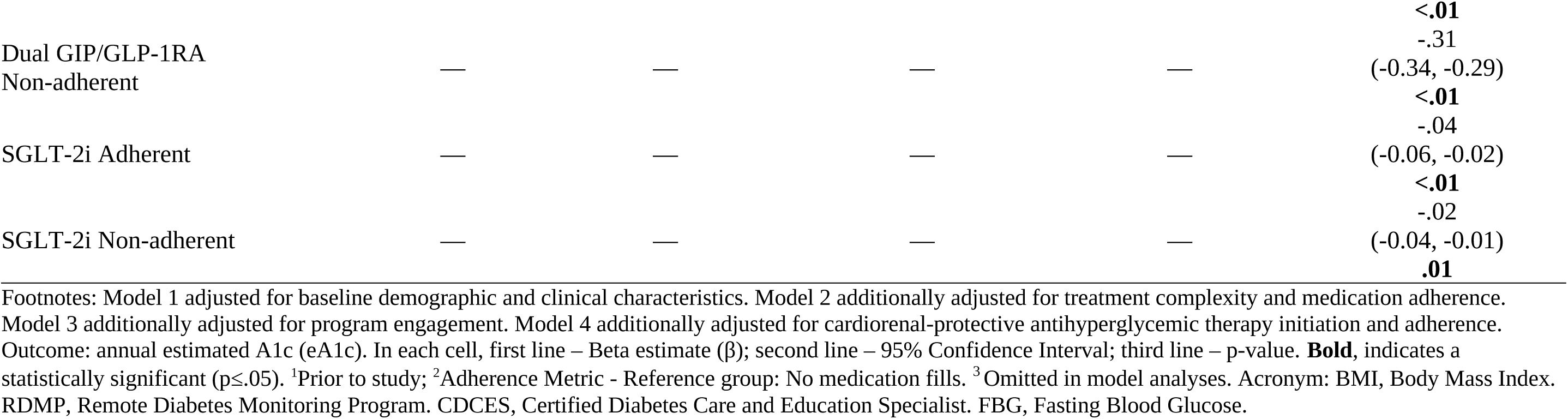
Longitudinal Mixed-Effects Models Examining Annual Trends in Estimated A1c.

In the pooled OLS models used for the Gelbach decomposition, the baseline-adjusted annual eA1c trend was −0.06 percentage points per year (95% CI, −0.07, −0.06), compared with −0.05 percentage points per year (95% CI; −0.06, −0.05) in the fully adjusted model. The total explained attenuation was −.010 percentage points per year (95% CI; −0.011, −0.010). The cardiorenal protective antihyperglycemics domain made the largest contribution to the difference from the base model that adjusted for demographics and baseline characteristics and the fully adjusted model (contribution, −0.02 percentage points per year (95% CI; −0.02, −0.01; Holm-adjusted p<.01). Its estimated contribution exceeded the attenuation because it was partially offset by the program utilization and engagement domain, which contributed to the opposite direction (0.007; 95% CI; 0.007, 0.008); Holm-adjusted p<.01) demonstrating a suppressor effect. Treatment complexity contributed approximately zero independently. Consequently, domain percentages were 172.9%, −73.1%, and 0.2%, respectively. (**Table 4**)

**Table 4.** Gelbach Decomposition of the Annual Estimated A1c Trend.

|  | Estimate eA1c % Points, by<br>Year (95% CI) | Unadjusted<br>p-value | Holm-Adjusted<br>p-value | Share of Explained<br>Attenuation |
| --- | --- | --- | --- | --- |
| Base annual trend (Model 1) | -0.07 (-0.07, -0.06) | <.01 | - | - |
| Fully adjusted annual trend (Model 4) | -0.06 (-0.06, -0.05) | <.01 | - | - |
| Total Explained Attenuation | -0.01 (-0.01, -0.02) | - | - | 100% |
| *Domains* |  |  |  |  |
| Treatment History and Complexity | -0.00 (-0.00, 0.00) | .94 | .94 | 1.1% |
| Antihyperglycemic with Cardio-Renal<br>Benefits Therapy Use | -0.02 (-0.02, -0.01) | <.01 | <.01 | 170.8% |
| Program Engagement | 0.01 (0.01, 0.01) | <.01 | <.01 | -71.9% |

Footnotes: Positive values indicate attenuation of the annual trend, whereas negative values indicate suppression effects that offset attenuation attributable to other domains. Model 1 (Demographics) vs Model 4 (Treatment Complexity, Program Engagement and Cardiorenal Protective Antihyperglycemic Adherence). Domain CIs and p-values are from member bootstrap B=2000, seed=20261005. Acronym: CI, Confidence interval. Gelbach: Pooled OLS with member-clustered.

### Secondary Outcome Analysis

The proportion of members with eA1c <7% increased throughout the study period. In the primary exchangeable-correlation GEE model, the unadjusted odds of attaining eA1c <7% increased by 10.1% per year (odds ratio [OR]=1.10; 95% CI 1.09, 1.11; p<.01), corresponding to an average marginal increase of 1.63 percentage points per year (95% CI 1.50, 1.75). After adjustment for demographics and baseline clinical characteristics and the three time-varying domains, the odds of attaining an eA1c <7% increased by 8% per year (OR=1.08; 95% CI 1.07, 1.09, p<.01), corresponding to an adjusted average marginal increase of 1.21 percentage points per year (95% CI; 1.08, 1.34). Model-based adjusted probabilities increased from 75.2% (95% CI 74.8%, 75.6%) in 2021 to 80.1% (95%CI; 79.8%, 80.5%). Results were consistent under independent and autoregressive working-correlation structures. The total explained attenuation was 0.14 (95% CI 0.08, 0.21) indicating that the time-varying domains explained 8.7% of the annual increase. Similar to the primary outcome analysis, cardiorenal protective antihyperglycemics were the largest contributor to the decomposition trend with program utilization and engagement demonstrating a suppressor effect and treatment complexity having no effect. The domain percentages were 242.8% (95% CI 179.2%, 374.0%), −154.1% (95% CI −281.0%, −103.9%), and 11.3% (95% CI; 34.9 (95% CI −34.9%, 57.4%), respectively. **(Tables 5a-c and 6)**

**Table 5a.**
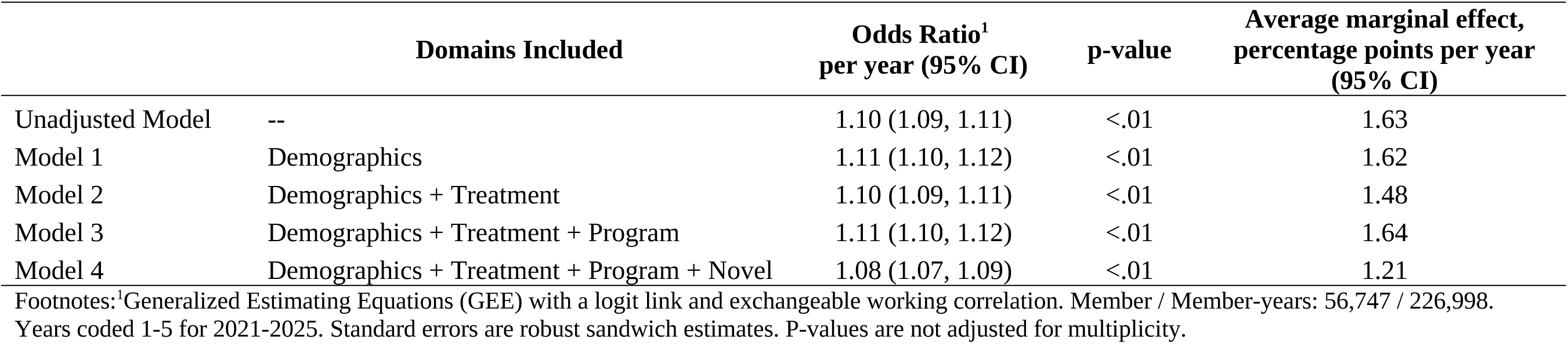
Annual Trend in Achievement of Estimated A1c <7%.

**Table 5b.** Predicted Probability (%) of Achieving Estimated A1c <7% by Calendar Year.

| Year | Unadjusted Model | Adjusted Full Model |
| --- | --- | --- |
| 2021 | 73.9 | 75.2 |
| 2022 | 75.7 | 76.5 |
| 2023 | 77.5 | 77.8 |
| 2024 | 79.1 | 79.0 |
| 2025 | 80.6 | 80.2 |

Footnote: <sup>1</sup>Predicted probabilities are average over each members observed covariate values (average predicted margins)

**Table 5c.** Sensitivity Analyses of the Model 4 Annual Trend Under Alternative Correlation Structures.

| Working correlation | Odds Ratio<br>per year (95% CI) | Average marginal effect, % points per year (95% CI) |
| --- | --- | --- |
| Exchangeable (primary) | 1.080 (1.071, 1.089) | 1.21 |
| Independence<sup>1</sup> | 1.101 (1.091, 1.110) | 1.49 |
| Autoregressive (AR-1) | 1.097 (1.087, 1.106) | 1.47 |

Footnote: <sup>1</sup>The independence structure is constant, even when time-varying covariates response to prior glycemic control

**Table 6.**
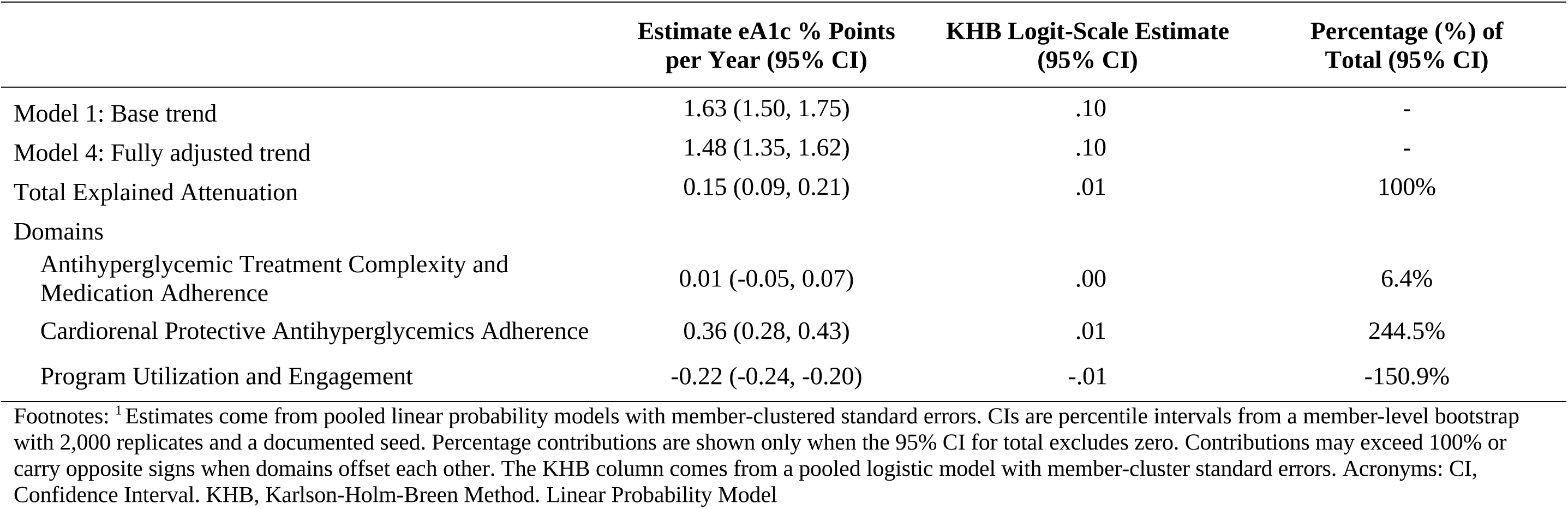
Karlson-Holm-Breen Method (KHB) Decomposition of Annual Trends in Achievement of Estimated A1c <7%. ^1^

### Sensitivity Analyses

Results were generally consistent with the prespecified analysis treating year as categorical. Before adjustment, mean eA1c was 0.214 percentage points lower in 2025 than in 2021 (95% CI, −0.222, −0.206). In the fully adjusted categorical year model, the 2025 versus 2021 contrast was 0.0007 percentage points (95% CI, −0.016, 0.029), indicating that the measured time-varying covariates accounted for the net difference between those years and that the intermediate annual estimates were not fully monotonic after complete adjustment, supporting cautious interpretation of calendar year as strictly a linear process. (**Supplemental Table 1**)

When analyses were stratified by self-reported A1c at enrollment, the fully adjusted annual decline was larger among members reporting an enrollment A1c ≥ 7% (β= −.05; 95% CI, −0.06, −0.04) than members reporting a value < 7% (β= −.03; 95% CI, −0.03, −0.02). Thus, as expected, members beginning above the conventional target experienced greater improvement, while those beginning below the target maintained glycemic control with smaller additional decline. (**Supplemental Tables 2aand 2b**).

## DISCUSSION

In this longitudinal cohort of adults with T2D who remained in the RDMP for 5 years, glycemic control was maintained and modestly improved. Adjusted mean eA1c fell by 0.18 percentage points from 2021 to 2025, while the adjusted probability of achieving eA1c < 7% increased from 75.2% to 80.1%. In the fully adjusted model, the odds of reaching that target rose 8% per year. These findings suggest durable glycemic control within this continuously observed cohort during a period of substantial adoption of cardiorenal protective antihyperglycemics.

The adoption and continued use of GLP-1RAs, dual GIP/GLP-1RAs, and SGLT-2is represent the largest measured contributions to changes in glycemic outcomes. In the primary Gelbach decomposition, this domain contributed more than the net explained attenuation of the eA1c trend because its association was partially offset by program utilization and engagement measures. Treatment complexity contributed little independently of the other two domains. A similar pattern was observed for the attainment of eA1c < 7%. The opposing contributions of the cardiorenal protective antihyperglycemics and program engagement measures illustrate that the observed five-year glycemic trajectory was associated with multiple concurrent changes rather than a single dominant factor. Because these results represent statistical deomposition of observational data, the domain contributions should not be interpreted as causal mediation effects.

Increasing evidence supports that GLP-1RAs and SGLT-2is target clinical outcomes beyond glycemic control demonstrating cardiovascular and renal benefits, while GLP-1RAs and dual GIP/GLP-1RAs also produce clinically meaningful weight loss.[4,10,11] This distinction is important when interpreting treatment adoption during the study period. Increasing use of these therapies may not have been driven solely by glycemic control need, but because their clinical benefits extend to risk reduction in other conditions. Consequently, impact to changes in eA1c during this period occurred within a broader evaluation of diabetes treatment in which medication selection increasingly reflects cardiometabolic and renal risks as well as glycemic control.

Program engagement showed a more complex relationship with glycemic control. More frequent FBG monitoring and greater web/mobile application use were associated with lower eA1c while having at least one coaching interaction was associated with slightly higher eA1c in the fully adjusted model. This association should not be interpreted as coaching worsens glycemic control. In the RDMP, coaching contact included outreach triggered by abnormal glucose readings, making greater coaching exposure potentially a marker of clinical or support needs that confound the program intervention with need.

The magnitude of the overall change in mean eA1c should also be interpreted in the context of the cohort’s relatively favorable glycemic control at the beginning of the study period with 75% of the members having an eA1c < 7% leaving comparatively limited opportunity for additional reduction in cohort-level mean eA1c. However, mean eA1c declined over the five years while the proportion of members achieving eA1c < 7% increased. Therefore, maintenance of glycemic control rather than magnitude of eA1c reduction is an important consideration when evaluating long-term diabetes management programs.

These findings should be considered alongside the growing literature on digital and virtual diabetes management programs. Meta-analyses of randomized trials have generally found that digital health and telemedicine interventions improve A1c compared with usual care, although effect sizes and interventions are heterogeneous. A recent meta-analysis of 118 randomized trials reported improvement in A1c with digital health interventions[12] while another analysis of 58 randomized trials estimated an average A1c reduction of approximately 0.38 percentage points relative to usual care.[13] However, much of the evidence evaluates relatively short intervention periods and heterogeneous combinations of telemedicine, mobile applications, education, and remote monitoring. This study adds a complementary perspective by evaluating glycemic trajectories over a recent five-year period within the same continuously observed cohort and by explicitly examining concomitant changes is pharmacotherapy and program utilization rather than attributing the longitudinal trajectory to the digital program alone.

This study has several limitations that should be considered. First, this is an observational study and residual and unmeasured confounding cannot be excluded. Second, the principal glycemic outcome was an estimated A1c derived from FBG values rather than laboratory A1c and therefore depends on members’ testing patterns which may vary over time. Medication exposure was derived from prescription-fill records, so adherence reflects medication availability rather than confirmed ingestion. Also, much of members’ medical care was directed by external providers limiting the ability to capture clinician recommendations related to medication management, glycemic monitoring, and diet/exercise recommendations that may influence glycemic control. In addition, lifestyle factors like diet quality and types of physical activity as well as social determinants of health could not be fully captured, especially as time-varying measures. Similarly, race and ethnicity were not systematically collected, so we could not assess whether glycemic trajectories, program engagement, or adoption of cardiorenal protective therapies varied across groups. We recognize this gap given documented disparities in SGLT-2is and GLP-1RAs.[14] Individuals with CGM use were excluded because the primary outcome depends on FBG measurements, which may limit generalizability as CGM utilization is being used more in the management of T2D.[15] Finally, requiring a repeated glycemic observation and prescription-fill information across all five study years selected a population with sustained program participation and data availability. Therefore, results should not be generalized to all individuals who participate in a RDMP.

These findings have implications for the design and evaluation of RDMPs and should inform future studies. First, the adoption of cardiorenal protective antihyperglycemics during this period addressed clinical outcomes and risks that extend beyond glucose control, so only evaluating glycemic status may capture just a portion of the benefit of these therapies when used with RDMPs. Therefore, RDMPs may increasingly benefit from a broader cardiometabolic framework inclusive of additional clinical outcome collection. Second, due to the confounding of program engagement with medication management and control status needs, clinically meaningful engagement metrics should be developed to assist in quantifying contribution of these engagements in improved outcomes. Third, future evaluations should incorporate standardized collection of race, ethnicity, and other sociodemographic characteristics to determine for whom RDMPs and concurrent pharmacotherapy are most effective, a gap highlighted in independent assessments of digital diabetes programs.[16]

## Conclusion

In this long-term experience with a large cohort of patients enrolled in a digital diabetes management program, progressive improvement in glycemic control was observed over five years of follow-up. The pattern of glycemic improvement was consistent regardless of background exposure to GLP1-RAs and SGLT-2is therapies, including in participants who were never exposed to these therapies. Because these therapies act across diabetes, cardiovascular, renal, and weight-related diseases, future program design should move from single-condition monitoring toward whole-person cardiometabolic care. These observations support guideline recommendations that include both the use of indicated medical therapies, as well as lifestyle modification which can be facilitated with digital remote programs.

## Author Declarations

All authors listed have seen and approved the manuscript.

## Distribution/Reuse Options

Distribution/Reuse Options: “All Rights Reserved: By selecting this option, you are retaining all rights over the use of your article. Anyone who wishes to share, reuse, remix, or adapt your article must first contact you for permission.”

## Data Availability

Data Availability Statement: “De-identified and aggregate data in this study are available from the corresponding author upon reasonable request, subject to institutional and ethical approvals.”

## ACKNOWLEDGEMENTS

The authors thank Dr. Marc Bonaca for his thoughtful discussions and insights that helped inform the development and refinement of the manuscript.

