## Supplemental Tables 1, 2a, and 2b for "Five-Year Glycemic Trends in a Remote Diabetes Monitoring Program (RDMP): Contributions of Program Engagement and Evolving Pharmacotherapy Adoption and Adherence"

**SUPPLEMENTARY TABLES: SENSITIVITY ANALYSIS RESULTS**

**Table S1. Longitudinal Mixed-Effects Models Examining Annual Trends in Estimated A1c, with Year as a categorical variable**

| | **Unadjusted Model** | **Model 1** | **Model 2** | **Model 3** | **Model 4** |
|---|---|---|---|---|---|
| | Year only (β, 95% CI, p-value) | Baseline Demographics and Clinical Characteristics | Treatment Complexity and Medication Adherence | Program Utilization and Engagement | Cardiorenal Protective Antihyperglycemics Adherence |
| **Year (Categorical)** | | | | | |
| 2021 (ref) | — | — | — | — | — |
| 2022 | -.02 (-.03, -.01) <.01 | -.02 (-.03, -.01) <.01 | -.02 (-.03, .00) .09 | .09 (.06, .11) <.01 | .16 (.14, .18) <.01 |
| 2023 | -.12 (-.13, -.11) <.01 | -.13 (-.14, -.12) <.01 | -.12 (-.14, -.11) <.01 | -.03 (-.05, .00) .02 | .06 (.04, .09) <.01 |
| 2024 | -.19 (-.20, -.18) <.01 | -.21 (-.22, -.20) <.01 | -.20 (-.21, -.18) <.01 | -.10 (-.12, -.08) <.01 | .02 (-.01, .04) .15 |
| 2025 | -.21 (-.22, -.21) <.01 | -.25 (-.26, -.24) <.01 | -.23 (-.25, -.22) <.01 | -.14 (-.16, -.12) <.01 | .01 (-.02, .03) .56 |
| ***Baseline Demographics and Clinical Characteristics*** | | | | | |
| **Age** | — | — | — | — | — |
| ≥65 | | -.41 (-.50, -.32) <.01 | -.41 (-.51, -.33) <.01 | -.40 (-.49, -.31) <.01 | -.42 (-.51, -.33) <.01 |
| 55–64 | | -.31 (-.40, -.22) <.01 | -.32 (-.40, -.23) <.01 | -.30 (-.39, -.21) <.01 | -.31 (-.40, -.23) <.01 |

| | **Unadjusted Model** | **Model 1** | **Model 2** | **Model 3** | **Model 4** |
|---|---|---|---|---|---|
| | Year only (β, 95% CI, p-value) | Baseline Demographics and Clinical Characteristics | Treatment Complexity and Medication Adherence | Program Utilization and Engagement | Cardiorenal Protective Antihyperglycemics Adherence |
| 40–54 | | -.17 (-.26, -.08) <.01 | -.17 (-.26, -.08) <.01 | -.16 (-.25, -.07) <.01 | -.17 (-.26, -.08) <.01 |
| 18–39 (ref) | | | | | |
| Sex, Female (ref: Male) | — | -.20 (-.23, -.18) <.01 | -.21 (-.23, -.19) <.01 | -.21 (-.23, -.19) <.01 | -.21 (-.23, -.18) <.01 |
| **Diabetes Duration (years)** | — | — | — | — | — |
| ≥15 years | — | -.08 (-.12, -.04) <.01 | -.08 (-.11, -.04) <.01 | -.06 (-.10, -.03) <.01 | -.06 (-.10, -.03) <.01 |
| 10–<15 years | — | -.03 (-.07, .01) .10 | -.03 (-.07, .01) .12 | -.02 (-.06, .02) .36 | -.02 (-.06, .02) .42 |
| 5–<10 years | — | -.01 (-.05, .02) .44 | -.01 (-.05, .02) .42 | .00 (-.04, .03) .87 | .00 (-.04, .04) .94 |
| <5 years | — | -.06 (-.09, -.03) <.01 | -.06 (-.10, -.03) <.01 | -.05 (-.08, -.02) <.01 | -.05 (-.08, -.02) <.01 |
| Not reported (ref) | — | — | — | — | — |
| Baseline A1c (self-report) | — | — | — | — | — |

| | **Unadjusted Model** | **Model 1** | **Model 2** | **Model 3** | **Model 4** |
|---|---|---|---|---|---|
| | Year only (β, 95% CI, p-value) | Baseline Demographics and Clinical Characteristics | Treatment Complexity and Medication Adherence | Program Utilization and Engagement | Cardiorenal Protective Antihyperglycemics Adherence |
| ≥ 7% | — | .16 (.13, .19) <.01 | .17 (.14, .20) <.01 | .17 (.14, .20) <.01 | .17 (.14, .21) <.01 |
| < 7% | — | -.26 (-.29, -.23) <.01 | -.26 (-.29, -.23) <.01 | -.25 (-.28, -.22) <.01 | -.25 (-.28, -.22) <.01 |
| Not reported (ref) | — | — | — | — | — |
| **BMI category** | — | — | — | — | — |
| Obesity Class 3 | — | .01 (-.07, .09) .85 | .01 (-.07, .10) .74 | .01 (-.07, .10) .72 | .03 (-.06, .11) .50 |
| Obesity Class 2 | — | -.02 (-.10, .06) .65 | -.01 (-.10, .07) .73 | -.01 (-.10, .07) .73 | -.01 (-.09, .08) .89 |
| Obesity Class 1 | — | -.05 (-.13, .03) .19 | -.05 (-.14, .03) .19 | -.05 (-.13, .03) .20 | -.05 (-.13, .03) .21 |
| Overweight | — | -.11 (-.19, -.03) <.01 | -.11 (-.19, -.03) <.01 | -.11 (-.19, -.03) <.01 | -.12 (-.20, -.04) <.01 |
| Normal | — | -.19 (-.28, -.11) <.01 | -.20 (-.28, -.11) <.01 | -.19 (-.28, -.11) <.01 | -.21 (-.29, -.12) <.01 |

| | Unadjusted Model | Model 1 | Model 2 | Model 3 | Model 4 |
|---|---|---|---|---|---|
| | Year only (β, 95% CI, p-value) | Baseline Demographics and Clinical Characteristics | Treatment Complexity and Medication Adherence | Program Utilization and Engagement | Cardiorenal Protective Antihyperglycemics Adherence |
| Underweight | — | -.19 (-.48, .10) .20 | -.19 (-.48, .10) .20 | -.20 (-.49, .09) .18 | -.22 (-.51, .07) .14 |
| Not reported (ref) | — | — | — | — | — |
| **Hypertension** | — | .00 (-.03, .04) .85 | .00 (-.03, .04) .82 | .01 (-.03, .04) .72 | .00 (-.03, .04) .80 |
| **RDMP Duration (years)**[1] | — | .03 (.02, .05) <.01 | .03 (.02, .05) <.01 | .03 (.02, .04) <.01 | .03 (.02, .04) <.01 |
| **Diabetes Drug Utilization**[1] | — | — | — | — | — |
| GLP-1RA | — | -.01 (-.04, .01) .35 | .03 (.00, .06) .06 | .03 (.00, .05) .07 | .10 (.07, .12) <.01 |
| Dual GIP/GLP-1RA[3] | — | omitted | omitted | omitted | omitted |
| SGLT-2i | — | .08 (.05, .11) <.01 | .11 (.08, .14) <.01 | .11 (.08, .14) <.01 | .13 (.10, .16) <.01 |
| DPP-4i | — | .11 (.08, .14) <.01 | .12 (.08, .15) <.01 | .11 (.08, .15) <.01 | .12 (.09, .15) <.01 |
| Sulfonylureas | — | .17 (.14, .19) <.01 | .20 (.17, .22) <.01 | .20 (.17, .22) <.01 | .19 (.16, .22) <.01 |

| | Unadjusted Model | Model 1 | Model 2 | Model 3 | Model 4 |
|---|---|---|---|---|---|
| | Year only (β, 95% CI, p-value) | Baseline Demographics and Clinical Characteristics | Treatment Complexity and Medication Adherence | Program Utilization and Engagement | Cardiorenal Protective Antihyperglycemics Adherence |
| Thiazolidinediones | — | -.04 (-.08, .01) .11 | .02 (-.02, .07) .34 | .02 (-.03, .07) .39 | .01 (-.04, .06) .68 |
| Meglitinides | — | omitted | omitted | omitted | omitted |
| Biguanides | — | .00 (-.03, .02) .76 | .03 (.01, .06) .01 | .03 (.01, .06) .02 | .02 (.00, .05) .10 |
| Combination Drugs | — | .06 (.03, .09) <.01 | .10 (.06, .14) <.01 | .10 (.06, .14) <.01 | .09 (.05, .12) <.01 |
| Insulin | — | .16 (.13, .19) <.01 | .22 (.18, .25) <.01 | .22 (.19, .26) <.01 | .22 (.19, .26) <.01 |
| ***Antihyperglycemic Treatment Complexity and Medication Adherence*** | | | | | |
| **Treatment Complexity** | — | — | — | — | — |
| Only insulin | — | — | .15 (.10, .21) <.01 | .15 (.10, .21) <.01 | .12 (.07, .18) <.01 |
| 2+ classes of antihyperglycemic medications, with insulin | — | — | -.17 (-.21, -.14) <.01 | -.17 (-.21, -.14) <.01 | -.08 (-.12, -.04) <.01 |
| 2+ classes of antihyperglycemic medications without insulin | — | — | -.13 (-.15, -.10) <.01 | -.12 (-.15, -.10) <.01 | -.01 (-.04, .02) .54 |

| | Unadjusted Model | Model 1 | Model 2 | Model 3 | Model 4 |
|---|---|---|---|---|---|
| | Year only (β, 95% CI, p-value) | Baseline Demographics and Clinical Characteristics | Treatment Complexity and Medication Adherence | Program Utilization and Engagement | Cardiorenal Protective Antihyperglycemics Adherence |
| 1 class of antihyperglycemic medications | — | — | -.02 (-.04, .01) .14 | -.01 (-.04, .01) .20 | .03 (.00, .05) .03 |
| No antihyperglycemic medications (ref) | — | — | — | — | — |
| **Medication Adherence[2]** | — | — | — | — | — |
| DPP-4 Adherent | — | — | .07 (.04, .10) <.01 | .07 (.05, .10) <.01 | .01 (-.01, .04) .27 |
| DPP-4 Non-adherent | — | — | .04 (.02, .07) <.01 | .04 (.02, .07) <.01 | .01 (-.01, .04) .30 |
| Sulfonylureas Adherent | — | — | .01 (-.01, .03) .22 | .02 (.00, .04) .04 | .00 (-.02, .02) .73 |
| Sulfonylureas Non-adherent | — | — | .03 (.01, .05) <.01 | .04 (.02, .06) <.01 | .03 (.01, .05) <.01 |
| Thiazolidinediones Adherent | — | — | -.09 (-.12, -.05) <.01 | -.08 (-.12, -.05) <.01 | -.09 (-.12, -.05) <.01 |
| Thiazolidinediones Non-adherent | — | — | -.03 (-.07, .00) .08 | -.03 (-.07, .00) .08 | -.04 (-.07, -.01) .03 |

| | **Unadjusted Model** | **Model 1** | **Model 2** | **Model 3** | **Model 4** |
|---|---|---|---|---|---|
| | Year only (β, 95% CI, p-value) | Baseline Demographics and Clinical Characteristics | Treatment Complexity and Medication Adherence | Program Utilization and Engagement | Cardiorenal Protective Antihyperglycemics Adherence |
| Biguanides Adherent | — | — | -.01 (-.02, .01) .52 | .00 (-.02, .01) .82 | -.03 (-.05, -.01) <.01 |
| Biguanides Non-adherent | — | — | .03 (.01, .04) <.01 | .03 (.01, .04) <.01 | .00 (-.02, .01) .65 |
| Combination Drug Adherent | — | — | .03 (.00, .06) .04 | .03 (.00, .06) .03 | -.01 (-.04, .03) .70 |
| Combination Drug Non-adherent | — | — | .01 (-.02, .04) .43 | .01 (-.02, .04) .50 | -.01 (-.04, .02) .45 |
| Any Insulin | — | — | -.09 (-.11, -.06) <.01 | -.06 (-.09, -.04) <.01 | -.03 (-.06, -.01) <.01 |
| ***Program Utilization and Engagement*** | | | | | |
| At least one contact with a CDCES Coach | — | — | — | .03 (.00, .05) .03 | .02 (.00, .04) .07 |
| Proportion of days with FBG check | — | — | — | -.14 (-.16, -.12) <.01 | -.13 (-.16, -.11) <.01 |
| Proportion of months with ≥1 web/mobile app login | — | — | — | -.10 (-.12, -.07) <.01 | -.09 (-.11, -.06) <.01 |

| | Unadjusted Model | Model 1 | Model 2 | Model 3 | Model 4 |
|---|---|---|---|---|---|
| | Year only (β, 95% CI, p-value) | Baseline Demographics and Clinical Characteristics | Treatment Complexity and Medication Adherence | Program Utilization and Engagement | Cardiorenal Protective Antihyperglycemics Adherence |
| Sharing of the Health Summary Report | — | — | — | .00 (.00, .00) .71 | .00 (.00, .00) .84 |
| ***Cardiorenal Protective Antihyperglycemics Adherence***[2] | | | | | |
| Adherent (PDC ≥ 0.8) | — | — | — | -.19 (-.21, -.17) | -.19 (-.20, -.17) <.01 |
| Non-adherent (at least one fill and PDC < 0.8) | — | — | — | -.14 (-.16, -.12) | -.14 (-.15, -.12) <.01 |
| Adherent (PDC ≥ 0.8) | — | — | — | -.47 (-.51, -.43) | -.52 (-.56, -.48) <.01 |
| Non-adherent (at least one fill and PDC < 0.8) | — | — | — | -.30 (-.32, -.27) | -.32 (-.34, -.29) <.01 |
| Adherent (PDC ≥ 0.8) | — | — | — | -.06 (-.08, -.04) | -.05 (-.07, -.03) <.01 |
| Non-adherent (at least one fill and PDC < 0.8) | — | — | — | -.04 (-.06, -.03) | -.04 (-.06, -.02) <.01 |
| **Model Fit Statistics** | | | | | |
| -2Log-likelihood | 708638.00 | 328375.30 | 327953.60 | 327726.10 | 326279.90 |
| AIC | 708652.00 | 328441.30 | 328049.60 | 327830.10 | 326395.90 |

| | **Unadjusted Model** | **Model 1** | **Model 2** | **Model 3** | **Model 4** |
|---|---|---|---|---|---|
| | Year only<br>(β,<br>95% CI,<br>p-value) | Baseline Demographics and Clinical Characteristics | Treatment Complexity and Medication Adherence | Program Utilization and Engagement | Cardiorenal Protective Antihyperglycemics Adherence |
| BIC | 708725.90 | 328764.40 | 328519.60 | 328339.20 | 326963.80 |

Footnotes: [1]Prior to study; [2]Adherence Metric - Reference group: No medication fills. [3] Omitted in model analyses. Model 1 adjusted for baseline demographic and clinical characteristics. Model 2 additionally adjusted for treatment complexity and medication adherence. Model 3 additionally adjusted for program engagement. Model 4 additionally adjusted for cardiorenal-protective antihyperglycemic therapy initiation and adherence. Outcome: annual estimated A1c (eA1c). In each cell, first line – Beta estimate (β); second line – 95% Confidence Interval; third line. Acronym: BMI, Body Mass Index. RDMP, Remote Diabetes Monitoring Program. CDCES, Certified Diabetes Care and Education Specialist. FBG, Fasting Blood Glucose.

**Table S2a. Longitudinal Mixed-Effects Models Examining Annual Trends in Estimated A1c, among members with self-reported A1c ≥7%**

| | **Unadjusted Model** | **Model 1** | **Model 2** | **Model 3** | **Model 4** |
|---|---|---|---|---|---|
| | Year only (β, 95% CI, p-value) | Baseline Demographics and Clinical Characteristics | Treatment Complexity and Medication Adherence | Program Utilization and Engagement | Cardiorenal Protective Antihyperglycemics Adherence |
| ***Baseline Demographics and Clinical Characteristics*** | | | | | |
| **Age** | | | | | |
| ≥65 | — | -.51 (-.67, -.35) <.01 | -.52 (-.68, -.36) <.01 | -.51 (-.67, -.35) <.01 | -.54 (-.70, -.38) <.01 |
| 55–64 | — | -.34 (-.50, -.19) <.01 | -.35 (-.50, -.19) <.01 | -.34 (-.50, -.19) <.01 | -.36 (-.51, -.20) <.01 |
| 40–54 | — | -.14 (-.30, .01) .07 | -.15 (-.31, .01) .06 | -.15 (-.30, .01) .07 | -.15 (-.31, .00) .05 |
| 18–39 (ref) | — | — | — | — | — |
| Sex, Female (ref: Male) | — | -.16 (-.20, -.12) <.01 | -.17 (-.20, -.13) <.01 | -.17 (-.21, -.13) <.01 | -.16 (-.20, -.12) <.01 |
| **Diabetes Duration (years)** | | | | | |
| ≥15 years | — | -.10 (-.17, -.04) <.01 | -.10 (-.17, -.04) <.01 | -.09 (-.15, -.03) <.01 | -.09 (-.15, -.03) <.01 |
| 10–<15 years | — | -.04 (-.10, .03) .32 | -.03 (-.10, .03) .33 | -.02 (-.09, .05) .53 | -.02 (-.09, .05) .55 |

| | **Unadjusted Model** | **Model 1** | **Model 2** | **Model 3** | **Model 4** |
|---|---|---|---|---|---|
| | Year only (β, 95% CI, p-value) | Baseline Demographics and Clinical Characteristics | Treatment Complexity and Medication Adherence | Program Utilization and Engagement | Cardiorenal Protective Antihyperglycemics Adherence |
| 5–<10 years | — | -.05 (-.11, .02) .18 | -.05 (-.11, .02) .17 | -.04 (-.10, .03) .30 | -.04 (-.10, .03) .31 |
| <5 years | — | -.13 (-.19, -.07) <.01 | -.14 (-.20, -.08) <.01 | -.12 (-.18, -.06) <.01 | -.12 (-.18, -.06) <.01 |
| Not reported (ref) | — | — | — | — | — |
| **BMI category** | | | | | |
| Obesity Class 3 | — | .11 (-.05, .27) .17 | .12 (-.04, .28) .15 | .12 (-.04, .28) .13 | .13 (-.03, .29) .11 |
| Obesity Class 2 | — | .09 (-.07, .25) .28 | .09 (-.07, .25) .26 | .10 (-.06, .26) .23 | .10 (-.06, .26) .22 |
| Obesity Class 1 | — | .01 (-.14, .17) .85 | .01 (-.14, .17) .86 | .02 (-.14, .17) .80 | .01 (-.14, .17) .85 |
| Overweight | — | -.06 (-.21, .10) .48 | -.06 (-.21, .10) .47 | -.05 (-.21, .10) .52 | -.06 (-.22, .09) .43 |
| Normal | — | -.14 (-.30, .03) .11 | -.14 (-.31, .02) .09 | -.14 (-.30, .03) .11 | -.16 (-.33, .01) .06 |

| | Unadjusted Model | Model 1 | Model 2 | Model 3 | Model 4 |
|---|---|---|---|---|---|
| | Year only (β, 95% CI, p-value) | Baseline Demographics and Clinical Characteristics | Treatment Complexity and Medication Adherence | Program Utilization and Engagement | Cardiorenal Protective Antihyperglycemics Adherence |
| Underweight | — | -.12 (-.69, .44) .67 | -.11 (-.68, .45) .69 | -.12 (-.69, .44) .67 | -.15 (-.71, .41) .60 |
| Not reported (ref) | — | — | — | — | — |
| Hypertension | — | -.03 (-.09, .03) .30 | -.03 (-.09, .03) .30 | -.03 (-.09, .03) .33 | -.03 (-.09, .03) .32 |
| RDMP Duration (years)[1] | — | .02 (.00, .04) .08 | .02 (.00, .04) .11 | .02 (-.01, .04) .16 | .02 (-.01, .04) .19 |
| Diabetes Drug Utilization[1] | | | | | |
| GLP-1RA | — | -.01 (-.06, .04) .68 | .03 (-.02, .07) .25 | .03 (-.01, .08) .19 | .09 (.04, .14) <.01 |
| Dual GIP/GLP-1RA[3] | — | omitted | omitted | omitted | omitted |
| SGLT-2i | — | .02 (-.02, .07) .35 | .05 (.01, .10) .02 | .06 (.01, .10) .01 | .07 (.02, .12) <.01 |
| DPP-4i | — | .06 (.01, .11) .01 | .07 (.02, .12) .01 | .07 (.01, .12) .01 | .08 (.02, .13) <.01 |
| Sulfonylureas | — | .11 (.07, .15) <.01 | .13 (.09, .17) <.01 | .13 (.08, .17) <.01 | .12 (.08, .17) <.01 |

| | Unadjusted Model | Model 1 | Model 2 | Model 3 | Model 4 |
|---|---|---|---|---|---|
| | Year only (β, 95% CI, p-value) | Baseline Demographics and Clinical Characteristics | Treatment Complexity and Medication Adherence | Program Utilization and Engagement | Cardiorenal Protective Antihyperglycemics Adherence |
| Thiazolidinediones | — | -.03 (-.11, .04) .36 | .02 (-.06, .10) .65 | .01 (-.06, .09) .72 | .01 (-.07, .09) .80 |
| Meglitinides | — | omitted | omitted | omitted | omitted |
| Biguanides | — | -.12 (-.17, -.07) <.01 | -.08 (-.13, -.03) <.01 | -.10 (-.15, -.05) <.01 | -.11 (-.16, -.06) <.01 |
| Combination Drugs | — | -.01 (-.07, .05) .69 | .02 (-.04, .09) .52 | .01 (-.05, .08) .69 | .01 (-.06, .07) .84 |
| Insulin | — | .10 (.06, .15) <.01 | .15 (.10, .20) <.01 | .15 (.10, .20) <.01 | .15 (.10, .20) <.01 |
| ***Antihyperglycemic Treatment Complexity and Medication Adherence*** | | | | | |
| **Treatment Complexity** | | | | | |
| Only insulin | — | — | .12 (.02, .21) .02 | .10 (.01, .20) .04 | .07 (-.02, .17) .13 |
| 2+ classes of antihyperglycemic medications, with insulin | — | — | -.21 (-.28, -.15) <.01 | -.25 (-.32, -.19) <.01 | -.18 (-.25, -.11) <.01 |
| 2+ classes of antihyperglycemic medications without insulin | — | — | -.17 (-.23, -.12) <.01 | -.20 (-.25, -.14) <.01 | -.12 (-.17, -.06) <.01 |

| | Unadjusted Model | Model 1 | Model 2 | Model 3 | Model 4 |
|---|---|---|---|---|---|
| | Year only (β, 95% CI, p-value) | Baseline Demographics and Clinical Characteristics | Treatment Complexity and Medication Adherence | Program Utilization and Engagement | Cardiorenal Protective Antihyperglycemics Adherence |
| 1 class of antihyperglycemic medications | — | — | -.05 (-.11, .00) .06 | -.07 (-.13, -.02) .01 | -.04 (-.10, .01) .13 |
| No antihyperglycemic medications (ref) | — | — | — | — | — |
| **Medication Adherence**[2] | | | | | |
| DPP-4 Adherent | — | — | .06 (.01, .11) <.01 | .07 (.03, .12) <.01 | .03 (-.02, .07) .28 |
| DPP-4 Non-adherent | — | — | .06 (.01, .10) .01 | .07 (.02, .11) <.01 | .04 (.00, .08) .07 |
| Sulfonylureas Adherent | — | — | .04 (.01, .07) .02 | .06 (.03, .09) <.01 | .04 (.01, .08) <.01 |
| Sulfonylureas Non-adherent | — | — | .04 (.01, .08) .01 | .06 (.03, .09) <.01 | .05 (.02, .09) <.01 |
| Thiazolidinediones Adherent | — | — | -.08 (-.14, -.02) <.01 | -.07 (-.13, -.01) .02 | -.07 (-.13, -.01) .02 |
| Thiazolidinediones Non-adherent | — | — | -.03 (-.09, .03) .30 | -.02 (-.08, .04) .45 | -.03 (-.09, .03) .28 |

| | **Unadjusted Model** | **Model 1** | **Model 2** | **Model 3** | **Model 4** |
|---|---|---|---|---|---|
| | Year only (β, 95% CI, p-value) | Baseline Demographics and Clinical Characteristics | Treatment Complexity and Medication Adherence | Program Utilization and Engagement | Cardiorenal Protective Antihyperglycemics Adherence |
| Biguanides Adherent | — | — | .01 (-.02, .03) .49 | .05 (.03, .08) <.01 | .04 (.02, .07) <.01 |
| Biguanides Non-adherent | — | — | .04 (.02, .07) <.01 | .08 (.05, .11) <.01 | .07 (.04, .10) <.01 |
| Combination Drug Adherent | — | — | .04 (-.01, .09) .12 | .08 (.03, .13) <.01 | .06 (.01, .11) .03 |
| Combination Drug Non-adherent | — | — | .04 (-.02, .09) .19 | .06 (.00, .11) .04 | .05 (-.01, .10) .09 |
| Any Insulin | — | — | -.07 (-.10, -.03) <.01 | -.03 (-.07, .01) .12 | -.01 (-.05, .02) .46 |
| ***Program Utilization and Engagement*** | | | | | |
| At least one contact with a CDCES Coach | — | — | — | .07 (.03, .11) <.01 | .07 (.03, .11) <.01 |
| Proportion of days with FBG check | — | — | — | -.10 (-.13, -.06) <.01 | -.07 (-.11, -.04) <.01 |
| Proportion of months with ≥1 web/mobile app login | — | — | — | -.11 (-.16, -.06) <.01 | -.10 (-.14, -.05) <.01 |

| | Unadjusted Model | Model 1 | Model 2 | Model 3 | Model 4 |
|---|---|---|---|---|---|
| | Year only<br>(β,<br>95% CI,<br>p-value) | Baseline Demographics and Clinical Characteristics | Treatment Complexity and Medication Adherence | Program Utilization and Engagement | Cardiorenal Protective Antihyperglycemics Adherence |
| Sharing of the Health Summary Report | — | — | — | .00<br>(.00, .00)<br>.39 | .00<br>(.00, .00)<br>.30 |
| ***Cardiorenal Protective Antihyperglycemics Adherence***[2] | | | | | |
| GLP-1RA Adherent | — | — | — | — | -.15<br>(-.18, -.12)<br><.01 |
| GLP-1RA Non-adherent | — | — | — | — | -.11<br>(-.13, -.08)<br><.01 |
| Dual GIP/GLP-1RA Adherent | — | — | — | — | -.51<br>(-.58, -.44)<br><.01 |
| Dual GIP/GLP-1RA Non-adherent | — | — | — | — | -.31<br>(-.36, -.27)<br><.01 |
| SGLT-2i Adherent | — | — | — | — | -.03<br>(-.06, .00)<br>.03 |
| SGLT-2i Non-adherent | — | — | — | — | -.01<br>(-.04, .01)<br>.34 |
| ***Model Fit Statistics*** | | | | | |
| -2Log-likelihood | 270645.50 | 132110.00 | 132020.60 | 131973.90 | 131603.10 |
| AIC | 270653.50 | 132166.00 | 132106.60 | 132067.90 | 131709.10 |
| BIC | 270691.50 | 132411.70 | 132484.10 | 132480.40 | 132174.30 |

| | Unadjusted Model | Model 1 | Model 2 | Model 3 | Model 4 |
|---|---|---|---|---|---|
| | Year only<br>(β,<br>95% CI,<br>p-value) | Baseline Demographics and Clinical Characteristics | Treatment Complexity and Medication Adherence | Program Utilization and Engagement | Cardiorenal Protective Antihyperglycemics Adherence |

Footnotes: [1]Prior to study; [2]Adherence Metric - Reference group: No medication fills. [3] Omitted in model analyses. Model 1 adjusted for baseline demographic and clinical characteristics. Model 2 additionally adjusted for treatment complexity and medication adherence. Model 3 additionally adjusted for program engagement. Model 4 additionally adjusted for cardiorenal-protective antihyperglycemic therapy initiation and adherence. Outcome: annual estimated A1c (eA1c). In each cell, first line – Beta estimate (β); second line – 95% Confidence Interval; third line. Acronym: BMI, Body Mass Index. RDMP, Remote Diabetes Monitoring Program. CDCES, Certified Diabetes Care and Education Specialist. FBG, Fasting Blood Glucose.

**Table S2b. Longitudinal Mixed-Effects Models Examining Annual Trends in Estimated A1c, among members with self-reported A1c <7%**

| | **Unadjusted Model** | **Model 1** | **Model 2** | **Model 3** | **Model 4** |
|---|---|---|---|---|---|
| | Year only (β, 95% CI, p-value) | Baseline Demographics and Clinical Characteristics | Treatment Complexity and Medication Adherence | Program Utilization and Engagement | Cardiorenal Protective Antihyperglycemics Adherence |
| ***Baseline Demographics and Clinical Characteristics*** | | | | | |
| **Age** | | | | | |
| ≥65 | — | -.33 (-.45, -.20) <.01 | -.34 (-.46, -.21) <.01 | -.33 (-.46, -.21) <.01 | -.35 (-.47, -.23) <.01 |
| 55–64 | — | -.27 (-.39, -.14) <.01 | -.27 (-.40, -.15) <.01 | -.27 (-.39, -.14) <.01 | -.28 (-.40, -.15) <.01 |
| 40–54 | — | -.20 (-.33, -.08) <.01 | -.20 (-.33, -.08) <.01 | -.20 (-.33, -.07) <.01 | -.20 (-.32, -.07) <.01 |
| 18–39 (ref) | — | — | — | — | — |
| Sex, Female (ref: Male) | — | -.23 (-.26, -.21) <.01 | -.24 (-.26, -.21) <.01 | -.24 (-.26, -.21) <.01 | -.23 (-.26, -.21) <.01 |
| **Diabetes Duration (years)** | | | | | |
| ≥15 years | — | -.06 (-.10, -.02) <.01 | -.06 (-.10, -.01) <.01 | -.04 (-.09, .00) .03 | -.05 (-.09, -.01) .02 |
| 10–<15 years | — | -.06 (-.10, -.01) .01 | -.06 (-.10, -.01) .02 | -.04 (-.09, .00) .06 | -.04 (-.09, .00) .06 |

| | **Unadjusted Model** | **Model 1** | **Model 2** | **Model 3** | **Model 4** |
|---|---|---|---|---|---|
| | Year only<br>(β,<br>95% CI,<br>p-value) | Baseline Demographics and Clinical Characteristics | Treatment Complexity and Medication Adherence | Program Utilization and Engagement | Cardiorenal Protective Antihyperglycemics Adherence |
| 5–<10 years | — | -.03<br>(-.07, .02)<br>.23 | -.03<br>(-.07, .02)<br>.24 | -.01<br>(-.06, .03)<br>.49 | -.01<br>(-.06, .03)<br>.47 |
| <5 years | — | -.03<br>(-.07, .01)<br>.10 | -.03<br>(-.07, .00)<br>.06 | -.02<br>(-.06, .01)<br>.22 | -.02<br>(-.06, .01)<br>.23 |
| Not reported (ref) | — | — | — | — | — |
| **BMI category** | | | | | |
| Obesity Class 3 | — | -.04<br>(-.15, .06)<br>.40 | -.04<br>(-.14, .07)<br>.50 | -.03<br>(-.14, .07)<br>.54 | -.02<br>(-.12, .09)<br>.73 |
| Obesity Class 2 | — | -.07<br>(-.17, .03)<br>.19 | -.06<br>(-.17, .04)<br>.25 | -.06<br>(-.16, .04)<br>.27 | -.05<br>(-.15, .06)<br>.37 |
| Obesity Class 1 | — | -.08<br>(-.18, .02)<br>.12 | -.08<br>(-.18, .02)<br>.13 | -.07<br>(-.18, .03)<br>.15 | -.07<br>(-.17, .03)<br>.17 |
| Overweight | — | -.12<br>(-.22, -.01)<br>.03 | -.12<br>(-.22, -.01)<br>.02 | -.11<br>(-.21, -.01)<br>.03 | -.12<br>(-.22, -.02)<br>.02 |
| Normal | — | -.20<br>(-.30, -.09)<br><.01 | -.20<br>(-.31, -.10)<br><.01 | -.20<br>(-.30, -.10)<br><.01 | -.21<br>(-.32, -.11)<br><.01 |

| | **Unadjusted Model** | **Model 1** | **Model 2** | **Model 3** | **Model 4** |
|---|---|---|---|---|---|
| | Year only (β, 95% CI, p-value) | Baseline Demographics and Clinical Characteristics | Treatment Complexity and Medication Adherence | Program Utilization and Engagement | Cardiorenal Protective Antihyperglycemics Adherence |
| Underweight | — | -.27 (-.60, .06) .10 | -.29 (-.62, .04) .09 | -.29 (-.62, .04) .09 | -.30 (-.63, .03) .07 |
| Not reported (ref) | — | — | — | — | — |
| Hypertension | — | .04 (.00, .07) .06 | .04 (.00, .08) .05 | .04 (.00, .08) .04 | .04 (.00, .08) .05 |
| RDMP Duration (years)[1] | — | .04 (.02, .06) <.01 | .04 (.02, .05) <.01 | .04 (.02, .05) <.01 | .04 (.02, .05) <.01 |
| Diabetes Drug Utilization[1] | | | | | |
| GLP-1RA | — | -.01 (-.04, .03) .73 | .05 (.01, .08) .01 | .05 (.01, .09) <.01 | .11 (.07, .15) <.01 |
| Dual GIP/GLP-1RA[3] | — | omitted | omitted | omitted | omitted |
| SGLT-2i | — | .12 (.08, .15) <.01 | .16 (.12, .20) <.01 | .16 (.12, .20) <.01 | .17 (.13, .21) <.01 |
| DPP-4i | — | .15 (.12, .19) <.01 | .15 (.11, .19) <.01 | .15 (.11, .19) <.01 | .15 (.11, .19) <.01 |
| Sulfonylureas | — | .17 (.14, .21) <.01 | .21 (.17, .24) <.01 | .20 (.17, .24) <.01 | .20 (.16, .23) <.01 |

| | Unadjusted Model | Model 1 | Model 2 | Model 3 | Model 4 |
|---|---|---|---|---|---|
| | Year only (β, 95% CI, p-value) | Baseline Demographics and Clinical Characteristics | Treatment Complexity and Medication Adherence | Program Utilization and Engagement | Cardiorenal Protective Antihyperglycemics Adherence |
| Thiazolidinediones | — | -.02 (-.08, .03) .42 | .03 (-.03, .09) .36 | .03 (-.03, .09) .37 | .02 (-.04, .08) .58 |
| Meglitinides | — | omitted | omitted | omitted | omitted |
| Biguanides | — | .05 (.02, .08) <.01 | .09 (.06, .12) <.01 | .08 (.05, .11) <.01 | .07 (.04, .10) <.01 |
| Combination Drugs | — | .11 (.06, .15) <.01 | .15 (.10, .20) <.01 | .14 (.09, .19) <.01 | .12 (.07, .17) <.01 |
| Insulin | — | .09 (.05, .13) <.01 | .13 (.09, .18) <.01 | .13 (.09, .18) <.01 | .13 (.08, .17) <.01 |
| ***Antihyperglycemic Treatment Complexity and Medication Adherence*** | | | | | |
| **Treatment Complexity** | | | | | |
| Only insulin | — | — | .02 (-.05, .09) .55 | .01 (-.06, .08) .75 | -.01 (-.08, .06) .69 |
| 2+ classes of antihyperglycemic medications, with insulin | — | — | -.25 (-.29, -.20) <.01 | -.27 (-.32, -.23) <.01 | -.20 (-.24, -.15) <.01 |
| 2+ classes of antihyperglycemic medications without insulin | — | — | -.18 (-.20, -.15) <.01 | -.19 (-.22, -.17) <.01 | -.09 (-.12, -.07) <.01 |

| | Unadjusted Model | Model 1 | Model 2 | Model 3 | Model 4 |
|---|---|---|---|---|---|
| | Year only (β, 95% CI, p-value) | Baseline Demographics and Clinical Characteristics | Treatment Complexity and Medication Adherence | Program Utilization and Engagement | Cardiorenal Protective Antihyperglycemics Adherence |
| 1 class of antihyperglycemic medications | — | — | -.05 (-.08, -.03) <.01 | -.07 (-.09, -.04) <.01 | -.03 (-.06, -.01) <.01 |
| No antihyperglycemic medications (ref) | — | — | — | — | — |
| **Medication Adherence**[2] | | | | | |
| DPP-4 Adherent | — | — | .11 (.07, .14) <.01 | .12 (.08, .15) <.01 | .07 (.04, .10) <.01 |
| DPP-4 Non-adherent | — | — | .08 (.05, .11) <.01 | .09 (.06, .12) <.01 | .06 (.03, .10) <.01 |
| Sulfonylureas Adherent | — | — | .03 (.00, .05) .04 | .04 (.01, .07) <.01 | .02 (.00, .05) .10 |
| Sulfonylureas Non-adherent | — | — | .04 (.01, .06) <.01 | .05 (.02, .07) <.01 | .04 (.01, .07) <.01 |
| Thiazolidinediones Adherent | — | — | -.05 (-.09, .00) .05 | -.04 (-.09, .01) .09 | -.04 (-.09, .00) .06 |
| Thiazolidinediones Non-adherent | — | — | .02 (-.03, .07) .40 | .03 (-.02, .07) .27 | .02 (-.02, .07) .34 |

| | Unadjusted Model | Model 1 | Model 2 | Model 3 | Model 4 |
|---|---|---|---|---|---|
| | Year only (β, 95% CI, p-value) | Baseline Demographics and Clinical Characteristics | Treatment Complexity and Medication Adherence | Program Utilization and Engagement | Cardiorenal Protective Antihyperglycemics Adherence |
| Biguanides Adherent | — | — | .02 (.01, .04) <.01 | .05 (.03, .07) <.01 | .03 (.01, .04) <.01 |
| Biguanides Non-adherent | — | — | .04 (.02, .05) <.01 | .06 (.04, .08) <.01 | .04 (.02, .06) <.01 |
| Combination Drug Adherent | — | — | .08 (.04, .12) <.01 | .10 (.06, .14) <.01 | .07 (.03, .11) <.01 |
| Combination Drug Non-adherent | — | — | .05 (.01, .09) <.01 | .07 (.03, .11) <.01 | .05 (.01, .09) .02 |
| Any Insulin | — | — | -.03 (-.06, .01) .10 | -.01 (-.04, .03) .70 | .03 (-.01, .06) .11 |
| ***Program Utilization and Engagement*** | | | | | |
| At least one contact with a CDCES Coach | — | — | — | .00 (-.03, .02) .78 | -.01 (-.03, .02) .60 |
| Proportion of days with FBG check | — | — | — | -.04 (-.06, -.02) <.01 | -.02 (-.04, .00) .05 |
| Proportion of months with ≥1 web/mobile app login | — | — | — | -.07 (-.10, -.05) <.01 | -.06 (-.09, -.03) <.01 |

| | **Unadjusted Model** | **Model 1** | **Model 2** | **Model 3** | **Model 4** |
|---|---|---|---|---|---|
| | Year only (β, 95% CI, p-value) | Baseline Demographics and Clinical Characteristics | Treatment Complexity and Medication Adherence | Program Utilization and Engagement | Cardiorenal Protective Antihyperglycemics Adherence |
| Sharing of the Health Summary Report | — | — | — | .00 (.00, .00) .08 | .00 (.00, .00) .07 |
| ***Cardiorenal Protective Antihyperglycemics Adherence***[2] | | | | | |
| GLP-1RA Adherent | — | — | — | — | -.15 (-.17, -.13) <.01 |
| GLP-1RA Non-adherent | — | — | — | — | -.12 (-.14, -.10) <.01 |
| Dual GIP/GLP-1RA Adherent | — | — | — | — | -.45 (-.50, -.40) <.01 |
| Dual GIP/GLP-1RA Non-adherent | — | — | — | — | -.28 (-.31, -.25) <.01 |
| SGLT-2i Adherent | — | — | — | — | -.03 (-.06, -.01) <.01 |
| SGLT-2i Non-adherent | — | — | — | — | -.03 (-.06, -.01) <.01 |
| ***Model Fit Statistics*** | | | | | |
| -2Log-likelihood | 267644.20 | 133372.00 | 133098.40 | 133060.20 | 132376.50 |
| AIC | 267652.20 | 133428.00 | 133184.40 | 133154.20 | 132482.50 |
| BIC | 267691.30 | 133681.90 | 133574.30 | 133580.30 | 132963.10 |

| | **Unadjusted Model** | **Model 1** | **Model 2** | **Model 3** | **Model 4** |
|---|---|---|---|---|---|
| | Year only<br>(β,<br>95% CI,<br>p-value) | Baseline Demographics and Clinical Characteristics | Treatment Complexity and Medication Adherence | Program Utilization and Engagement | Cardiorenal Protective Antihyperglycemics Adherence |

Footnotes: [1]Prior to study; [2]Adherence Metric - Reference group: No medication fills. [3] Omitted in model analyses. Model 1 adjusted for baseline demographic and clinical characteristics. Model 2 additionally adjusted for treatment complexity and medication adherence. Model 3 additionally adjusted for program engagement. Model 4 additionally adjusted for cardiorenal-protective antihyperglycemic therapy initiation and adherence. Outcome: annual estimated A1c (eA1c). In each cell, first line – Beta estimate (β); second line – 95% Confidence Interval; third line. Acronym: BMI, Body Mass Index. RDMP, Remote Diabetes Monitoring Program. CDCES, Certified Diabetes Care and Education Specialist. FBG, Fasting Blood Glucose.
